# Regional [^18^F]RO948 tau-PET across the AD continuum in relation to plasma biomarkers, cognition and atrophy

**DOI:** 10.64898/2025.12.17.25342292

**Authors:** Mariola Zapater-Fajari, Marco Bucci, Konstantinos Chiotis, Ove Almkvist, Anders Wall, Jonas Eriksson, Gunnar Antoni, Ilaria Pola, Kübra Tan, Wiebke Traichel, Andrea L. Benedet, Nicholas J. Ashton, Kaj Blennow, Henrik Zetterberg, Nenad Bogdanovic, Agneta Nordberg

## Abstract

Understanding tau pathology progression across the Alzheimer’s disease (AD) continuum is critical for diagnosis and stratification. This study examined how age of onset and disease stage influence regional tau deposition using [¹⁸F]RO948-PET, and its relationship with plasma biomarkers, cognition, and cortical atrophy. In total, 57 participants underwent tau-PET, MRI, blood sampling, and neuropsychological testing: 39 patients with MCI (Aβ−/Aβ+) or AD, and 18 cognitively normal controls. The MCI Aβ+ and AD groups were further divided into early-onset (EOAD, <65y) and late-onset (LOAD, >65y) subgroups. MCI Aβ+ patients showed early tau accumulation in medial-temporal regions, extending to inferior-temporal cortex. MCI-EOAD exhibited more advanced neocortical tau binding, while MCI-LOAD showed intermediate lateral temporal involvement. In AD, EOAD patients had higher parietal tau burden than LOAD. Plasma biomarkers (p-tau181, p-tau217, p-tau231, GFAP, NFL) were elevated in MCI Aβ+ and AD. Plasma p-tau217 showed strong correlations with tau-PET in medial and inferior temporal regions, with weaker correlations in neocortical areas. EOAD showed non-linear tau-PET/p-tau217 associations, contrasting with LOAD’s linear pattern. Tau-PET was negatively correlated with global cognition and executive function, while p-tau217 better reflected early episodic memory decline. Both tau measures correlated with cortical thinning, especially in the entorhinal cortex. These findings highlight [¹⁸F]RO948-PET’s sensitivity in detecting early tau pathology and superiority in capturing individual differences in tau burden, particularly in advanced stages where plasma biomarkers plateaued. Tau-PET demonstrated superior resolution of disease progression and individual variability, reinforcing its value as a prognostic biomarker and a critical tool for patient stratification in clinical trials.

**One Sentence Summary:** [¹⁸F]RO948 tau-PET detects early tau pathology and onset-related patterns, outperforming plasma biomarkers in tracking Alzheimer’s progression.

## INTRODUCTION

Alzheimer’s disease (AD) is a progressive neurodegenerative disorder characterized by multiple pathological events occurring during decade(s) of disease progression. Accumulation of amyloid-β (Aβ) plaques and hyperphosphorylated tau neurofibrillary tangles have been considered key pathological markers of AD but recent data underline the importance of early inflammatory changes and synaptic dysfunction as well as changes in fluid biomarkers.(*1*) Gradual accumulation of amyloid-β (Aβ) in the brain is widely considered the initiating event in the AD pathological cascade, acting as a driving factor that facilitates tau pathology, which in turn shows stronger associations with neurodegeneration and clinical symptoms.(*2*) Elucidation of the in vivo mechanisms underlying these pathological processes has been substantially advanced through developments in molecular neuroimaging techniques. Positron emission tomography (PET) allows in vivo characterization of tau pathology, providing critical insights into regional tau brain deposition and its role in AD progression. Tau-PET imaging, with the first-generation tracers, such as [^18^F]AV-1451 (Flortaucipir), has revealed a stereotypical spread of tau pathology from the medial temporal lobe to neocortical regions,(*2–4*) aligning with Braak staging assessed in neuropathological studies.(*5, 6*) Nevertheless, considerable heterogeneity exists in tau accumulation across individuals, influenced by factors such as age at onset of the disease, genetic background, and sex, which reflect differences in clinical symptoms.(*7–9*) Consequently, tau accumulation patterns, as determined by PET, closely mirrors clinical symptoms, with progressive increases in tau-PET tracer binding correlating with disease progression(*10*) and predicting subsequent brain atrophy.(*11–16*) However, tau-PET first-generation tracers are affected by off-target binding in important regions like the choroid plexus (particularly compromising evaluation of tracer uptake in the medial temporal lobe, one of the earliest regions to show tau pathology).(*16–18*) The development of second-generation tau-PET tracers, such as [^18^F]MK6240, [^18^F]RO948, [^18^F]PI2620 offers improved specificity and lower non-specific binding, making them particularly suitable for studying tau deposition in the early stages of the disease and for differentiating AD from other neurodegenerative dementias.(*19*)

Tau-PET imaging offers high specificity, but its limitations in accessibility and routine clinical use highlight the importance of developing complementary biomarkers that are less invasive and more broadly applicable. Recent studies have shown the potential usefulness of different plasma biomarkers including tau phosphorylated at amino acids 217, 231 or 181 (p-tau217, p-tau231, p-tau181), glial fibrillary acidic protein (GFAP) and neurofilament light chain (NFL). Specifically, the p-tau217 variant has been demonstrated as a reliable and scalable biomarker for AD, offering potential utility for early detection and disease monitoring.(*20, 21*) Although variability in p-tau217 levels seems to be explained by both amyloid and tau pathology, elevated levels tend to emerge almost exclusively in individuals who are amyloid-positive.(*22, 23*) Before plasma biomarkers can be implemented in clinical settings, it is of highly relevance and importance to investigate their relationship with the more established imaging and CSF biomarkers of AD. Integrating tau-PET and MRI imaging with fluid biomarkers in a clinical cohort offers a promising approach to better understand the biological heterogeneity of AD and may shed light on more personalized diagnostic and therapeutic strategies.(*24*)

The present study aimed to provide a deeper understanding of the regional brain spreading of tau across the AD continuum using the second-generation tau-PET tracer [^18^F]RO948 and explore its relationships with plasma biomarkers, atrophy (MRI cortical thickness and volumes) and cognition. Our mission is to advance our understanding of interacting pathological mechanisms across the AD continuum, thereby enabling improved participant selection and evaluation in clinical trials, as well as the development of optimal disease-modifying treatments within a personalized medicine framework.

## MATERIALS AND METHODS

### Study design and Participants

This study consisted of 57 participants of which 39 were recruited from the Cognitive Assessment Unit at Karolinska University Hospital Huddinge, Stockholm, Sweden, to which they initially had been referred for assessments due to cognitive problems and 18 cognitively normal individuals (10 recruited from the Clinical Trial Consultants AB (Uppsala, Sweden), and 8 participant (non-carriers) from a longitudinal observational study of familial autosomal dominant AD at Karolinska Institutet. At the Cognitive Assessment Unit, patients underwent an extensive clinical evaluation including comprehensive history, physical and neurological exam, cognitive assessment, CSF sampling for both total tau (CSF t-tau), phosphorylated tau (CSF p-tau181), and amyloid-beta (CSF Aβ40 and Aβ42), CT or MRI, and for some cases [^18^F]FDG-PET and amyloid-PET ([^18^F]flutemetamol or [^11^C]PIB). Consensus diagnosis was established by a multidisciplinary dementia expert team comprising specialists in cognitive disorders, clinical neuropsychologists, and specialist nurses. The patients were diagnosed as Mild Cognitive Impairment (MCI) Aβ-, MCI Aβ+ and AD. Furthermore, MCI Aβ+ and AD groups were further classified according to age at-onset of the disease (65 years cut-off) as MCI Aβ+ Early-Onset (MCI-EOAD) and MCI Aβ+ Late-Onset (MCI-LOAD), and AD Early-Onset (EOAD) and AD Late-Onset (LOAD). During the experimental day, all 57 participants underwent, brain MRI, tau-PET [^18^F]RO948 and plasma sampling at the PET Center at Uppsala University (Uppsala, Sweden). All participants gave written informed consent to participate in the study, which was conducted according to the Declaration of Helsinki. The study protocol was approved by the Ethics review authority of Sweden.

### PET and MRI Image acquisition and processing

Dynamic [^18^F]RO948 PET scans were acquired on a GE Discovery^TM^ MI PET/CT system (General Electric Medical Systems, USA) starting 70–90 min after injection, divided into 4 frames of 300 seconds each. Participants received an intravenous dose of 3 MBq/kg (minimum 150 MBq, maximum 300 MBq). Images were reconstructed using the VPFX-S algorithm with three iterations and thirty-four subsets, including a standard Z-axis filter and a 3 mm post-reconstruction Gaussian filter (Matrix 256 × 256 pixels, field of view (FoV) 25 cm).

The 3D T1-weighted magnetic resonance imaging (MRI) scans were acquired in a SIGNA PET/MR 3.0 T MRI scanner (General Electric Medical Systems, USA), using a 3D inversion recovery fast spoiled gradient echo sequence (IR-FSPGR; GE BRAVO equivalent) with 1-mm isotropic voxel resolution (TR=8.49, TE=3.25, TI=450ms, flip=12**°)**. Three patients were scanned at Karolinska University Hospital, Huddinge due to having a pacemaker, using GE Discovery MR750w 3 T (BRAVO ART; 1 mm isotropic, TR = 11.2 ms, TE = 4.7 ms, TI = 450 ms, flip = 12°); Siemens MAGNETOM Skyra 3 T (GRE Dixon; 2 mm, TR = 5.9 ms, TE = 2.46 ms, flip = 12°); or Siemens MAGNETOM Sola Fit 1.5 T (MPRAGE; 1 mm, TR = 2200 ms, TE = 3.0 ms, TI = 900 ms, flip = 8°) MRI scanners. For one participant without an MRI scan, a CT image was used. The convolutional neural network SynthSR was applied to generate a validated synthetic MPRAGE-equivalent image from the CT.(*62*)

The resulting PET images were motion corrected, summed and co-registered to their corresponding T1-weighted MR images using SPM12 (Wellcome Department of Cognitive Neurology, London, UK; http://www.fil.ion.ucl.ac.uk/spm) and an in-house adaptation of the medical imaging pipeline MAGIA (https://aivo.utu.fi/magia/).(*63*) The inferior cerebellar cortex was used as reference region using the SUIT atlas to create standardized uptake value ratio (SUVR) images.(*17, 27, 64*) FreeSurfer (v7.4.1) was used to generate cortical parcellations based on the Desikan-Killiany atlas(*65*) and subcortical volumetric segmentations from T1-weighted MRI scans. These were applied to the tau-PET scans to extract mean regional SUVR values for each participant using the same in-house pipeline. For the main analyses, we selected a subset of FreeSurfer-derived ROIs, based on previous studies highlighting both regional heterogeneity in early tau deposition,(*2, 3*) and age at onset differences in regional tau accumulation.(*9, 66*) Key regions included the inferior temporal, middle temporal and superior temporal cortex, entorhinal cortex, hippocampus and amygdala were used as defined by the Desikan-Killiany atlas. In addition, three composite ROIs were created by grouping anatomically related regions including the frontal, parietal, and occipital cortices, as previously described.(*67*) For simplicity and consistency, all ROIs were averaged across hemispheres. For voxelwise analyses, parametric SUVR PET images were spatially transformed to the common Montreal Neurological Institute (MNI152) Space. Cortical thickness in FreeSurfer-derived ROIs and composites were computed as the distance from the gray matter/white matter boundary to the corresponding pial surface.(*68*) Hippocampal and amygdala volumes were extracted from the Freesurfer automated volumetric segmentation and divided by the estimated Total Intracranial Volume (TIV).(*69*)

### Plasma biomarkers

Blood samples were collected by venipuncture, into sodium-heparin Vacutainer® tubes (BD Diagnostics). Samples were centrifuged for 10 min at 1500 g at 4 °C, within an hour of sampling, and the supernatant plasma was aliquoted into 1 ml polypropylene tubes and frozen at -80 °C.

Plasma samples (of p-tau217, p-tau181, p-tau231, Aβ40, Aβ42, GFAP, NFL and APOE4) were measured using the NULISAseq (Alamar) CNS panel(*70*) according to the manufacturer’s instructions. NULISA biomarkers are expressed in NULISA Protein Quantification (NPQ) units.

### Cognitive assessment

All individuals underwent extensive cognitive assessment. Briefly, six tests were used to derive four different cognitive domains. Verbal episodic memory was measured using the Rey Auditory Verbal Learning Test (RAVLT) and the free recall score after 30 minutes (RAVLT retention) tests.(*71*) The Rey-Osterrieth Complex Figure delayed recall (RO retention) was used to assess visuospatial episodic memory.(*71*) Executive function was assessed with both the Trail Making Test B (TMT-B),(*71*) and the Coding subtest from the Wechsler Adult Intelligence Scale – Fourth Edition (WAIS-IV).(*72*) Finally, short term memory/attention was measured using Trail Making Test A (TMT-A).(*71*) Participant neuropsychological testing scores were converted to z-scores using the CN group as reference. Composite scores for each cognitive domain were calculated by averaging the z-scores of the corresponding individual tests. Lower (i.e. more negative) z-scores were representative of greater cognitive impairment. We used Mini-Mental State Examination (MMSE)(*73*) as a global cognitive measure. Mean time from neuropsychological testing to biomarker testing was 1.30 years (SD=2.13).

### Statistical analyses

All analyses were carried out in R version 4.4.3 (https://www.r-project.org). Data visualization was conducted using *ggplot2* package, and regional brain maps were generated with *ggseg*. Participants with a non-AD diagnosis (MCI Aβ-) were excluded from the main analyses to ensure a homogeneous AD-focused sample.

Group comparisons were performed using the Kruskal-Wallis test, with Wilcoxon rank-sum post hoc test for pairwise comparisons.

Following a similar procedure described in Cho et al.(*3*) regional tau involvement was determined first by computing z-scores for each participant using the CN individuals as reference. Regions with z-scores>2 were classified as tau-positive. For each region, the proportion of participants showing involvement was calculated. Regions with higher frequency of tau positivity are interpreted as more commonly and/or earlier affected in the disease course.

For a more detailed fine-grained differences in [^18^F]RO948 tau-PET signal voxel-wise group comparisons were performed with SPM12 using two sample t-tests. Only clusters surviving family-wise error (FWE) correction at p < 0.05 at the cluster level were considered statistically significant. First, we compared MCI Aβ-, MCI Aβ+, and dementia AD groups against CN participants. Then, subgroup comparisons were performed between EOAD and LOAD within both MCI and AD stages. Results were visualized using BrainNet Viewer with interpolated surface mapping.(*74*)

Spearman correlations were performed with pairwise deletion. To test [^18^F]RO948 tau-PET age changes in a continuous manner, we first evaluated the relationship between [^18^F]RO948 tau-PET and age for the entire sample and within groups of CN and AD continuum (MCI Aβ+ and AD). To assess whether the relationship between [^18^F]RO948 tau-PET and plasma biomarkers differed by age at onset group (EOAD vs. LOAD), we employed Generalized Additive Models (GAMs) including an interaction term (plasma × age group).(*75*) GAMs were implemented using the *mgcv* package (method = “REML”, family = Gamma (link = “log”)). To better model the continuum of aging, younger CN individuals (age<65) were included in the EOAD group curve, and older CN participants (age>65) were included in the LOAD group curve. To directly test whether the smooth functions differed between age at onset groups, Wald tests were conducted on the differences of the estimated smooth term coefficients. This approach provides a targeted statistical test of the interaction effect on the shape of the plasma-tau-PET relationship.(*76*) GAMs were also used as a visualization tool to explore potential non-linear associations between variables of interest, using smooth functions without imposing *a priori* assumptions on the exact form of the relationship. To test whether tau PET or plasma biomarkers differentially predicted cognitive outcomes or cortical thickness or volumes in EOAD and LOAD groups, we initially explored non-linear relationships using GAMs but given the linear nature of the associations and better interpretability, linear models were ultimately used. Interaction terms (e.g., region or plasma biomarker × age group) were included to examine potential differences between EOAD and LOAD. Sensitivity analyses were conducted to examine potential sex differences in all relationships. In addition, all analyses were adjusted for age, sex, and *APOE* status.

To control multiple comparisons in pairwise correlation analyses, false discovery rate (FDR) correction (Benjamini-Hochberg) was applied to the resulting *p*-values. Adjusted *p*-values < 0.05 were considered statistically significant.

## RESULTS

### Participant characteristics

Table 1 presents all 57 participants which consisted of 18 cognitive healthy controls, 10 MCI Aβ-, 16 MCI Aβ+ (9 MCI-EOAD and 7 MCI-LOAD), 13 AD patients (5 EOAD and 8 LOAD). Table 1 presents the general characteristics of the individuals, including demographic, clinical, and biomarker data across diagnostic groups. Additional biomarker and cognitive data are also presented in Table S1. The Aβ+ groups (MCI and AD) presented higher *APOE* ε4 carriership compared with CN and MCI Aβ- individuals (Table 1). Moreover, they presented the worst cognitive performance and had lower cortical thickness than CN individuals, with significant differences varying across regions and domains. Although not statistically significant, EOAD individuals showed poorer cognitive performance in episodic memory and visuospatial memory than LOAD but similar volumetric or thickness measures (Table 1 and Table S1).

**Table 1.** Participant characteristics. Values are presented as Mean (SD) (continuous variables) or n (%) (categorical variables). Shown are demographic variables, APOE ε4 carriership, cerebrospinal fluid (CSF) and plasma phosphorylated tau (p-tau217), tau-PET regional Standardized Uptake Value Ratio (SUVR), cortical thickness measures, and cognitive z-scores across cognitively normal (CN), mild cognitive impairment amyloid-negative (MCI Aβ–), mild cognitive impairment amyloid-positive (MCI Aβ+), and Alzheimer’s disease (AD) groups, for both early-onset (EOAD) and late-onset (LOAD) cases. Group comparisons displayed: CN vs MCI AB+ vs AD, MCI-EOAD vs MCI-LOAD, EOAD vs LOAD. a = statistically different from CN, b = statistically different from MCI AB+. c = statistically different from MCI-EOAD, d = statistically different from EOAD. (*p*<0.05) Pathological thresholds for CSF-pTau181: 56.6 pg/ml and for CSF Aβ42/40: 0.68 pg/ml. MMSE= Mini-Mental State Examination.

| Variable | CN N<br>= 18 | MCI AB-<br>N = 10 | MCI AB+ N<br>= 16 | AD N<br>= 13 | MCI-<br>EOAD<br>N = 9 | EOAD<br>N = 5 | MCI-<br>LOAD<br>N = 7 | LOAD<br>N = 8 |
| --- | --- | --- | --- | --- | --- | --- | --- | --- |
| Age (years) | 65 (9) | 78 (3) | 68 (8) | 71 (10) | 61 (5) | 61 (3) | 75 (3)<sup>c</sup> | 77 (6)<sup>d</sup> |
| Sex (%) |  |  |  |  |  |  |  |  |
| Female | 8 (44%) | 3 (30%) | 9 (56%) | 6 (46%) | 5 (56%) | 2<br>(40%) | 2 (29%) | 5 (63%) |
| Male | 10<br>(56%) | 7 (70%) | 7 (44%) | 7 (54%) | 4 (44%) | 3<br>(60%) | 5 (71%) | 3 (38%) |
| Education (years) | 12.59<br>(3.00) | 15.00<br>(2.16) | 15.21<br>(1.47)<sup>a</sup> | 13.63<br>(2.45) | 14.17<br>(1.61) | 12.33<br>(0.58) | 16.00<br>(0.82) | 14.40<br>(2.88) |
| MMSE | 28.61<br>(1.38) | 27.33<br>(1.58) | 26.00 (4.08) | 24.46<br>(3.07)<sup>a</sup> | 26.00<br>(3.46) | 23.60<br>(2.07) | 26.00<br>(4.90) | 25.00<br>(3.59) |
| APOE E4 carriership<br>(%) |  |  |  |  |  |  |  |  |
| E4 | 4 (22%) | 2 (20%) | 13 (81%)<sup>a</sup> | 11 (85%)<sup>a</sup> | 7 (78%) | 5<br>(100%) | 6 (86%) | 6 (75%) |
| CSF p-tau181 |  | 48 (17) | 79 (45) | 112 (68) | 70 (47) | 119<br>(67) | 90 (44) | 107<br>(73) |
| CSF Aβ42/40 |  | 0.99<br>(0.17) | 0.54 (0.12) | 0.47 (0.06) | 0.57<br>(0.12) | 0.45<br>(0.06) | 0.51<br>(0.12) | 0.49<br>(0.06) |
| MTA |  |  |  |  |  |  |  |  |
| 0-1 |  | 5 (50%) | 10 (63%) | 8 (62%) | 7 (78%) | 3<br>(60%) | 3 (43%) | 5 (63%) |
| 2 |  | 1 (10%) | 6 (38%) | 4 (31%) | 2 (22%) | 2<br>(40%) | 4 (57%) | 2 (25%) |
| 3-4 |  | 4 (40%) | 0 (0%) | 1 (7.7%) | 0 (0%) | 0 (0%) | 0 (0%) | 1 (13%) |
| Plasma p-tau217 | 11.03<br>(0.39) | 11.60<br>(0.32) | 12.10<br>(0.45)<sup>a</sup> | 12.66<br>(0.38)<sup>ab</sup> | 11.94<br>(0.52) | 12.55<br>(0.34) | 12.31<br>(0.22) | 12.72<br>(0.41) |
| Entorhinal Cortex<br>SUVR | 1.23<br>(0.14) | 1.31<br>(0.19) | 1.91 (0.42)<sup>a</sup> | 2.21 (0.41)<sup>a</sup> | 1.91<br>(0.54) | 2.07<br>(0.52) | 1.92<br>(0.24) | 2.29<br>(0.34) |
| Inferior Temporal<br>Cortex SUVR | 1.35<br>(0.15) | 1.32<br>(0.12) | 1.83 (0.65)<sup>a</sup> | 2.29 (0.71)<sup>a</sup> | 1.90<br>(0.80) | 2.41<br>(0.96) | 1.75<br>(0.44) | 2.22<br>(0.58) |
| Parietal Cortex SUVR | 1.19<br>(0.12) | 1.16<br>(0.14) | 1.41 (0.48) | 1.84<br>(0.66)<sup>ab</sup> | 1.58<br>(0.59) | 2.39<br>(0.72) | 1.19<br>(0.09) | 1.50<br>(0.34) |
| Attention z-score | 0.00<br>(1.00) | -3.80<br>(2.23) | -3.52 (2.16)<sup>a</sup> | -3.33 (2.03) | -4.18<br>(2.55) | -2.05<br>(2.21) | -2.43<br>(0.66) | -3.96<br>(1.91) |
| Episodic Memory z-score | 0.00<br>(0.95) | -1.22<br>(0.70) | -1.26 (1.64)<sup>a</sup> | -1.58 (0.78) | -1.30<br>(1.48) | -2.26<br>(0.19) | -1.19<br>(2.06) | -0.91<br>(0.42) |
| Visuospatial Memory<br>z-score | 0.00<br>(1.00) | -1.32<br>(1.21) | -0.92 (1.37) | -1.67 (0.98) | -1.23<br>(1.19) | -2.11<br>(1.15) | -0.49<br>(1.61) | -1.34<br>(0.84) |
| Executive Function z-score | 0.03<br>(0.85) | -1.72<br>(1.24) | -1.46 (1.73)<sup>a</sup> | -1.90 (1.27) | -1.57<br>(2.06) | -1.70<br>(1.83) | -1.28<br>(1.23) | -2.07<br>(0.80) |
| Entorhinal Cortex<br>Thickness | 3.32<br>(0.26) | 2.81<br>(0.31) | 2.96 (0.29)<sup>a</sup> | 2.73 (0.34) | 3.00<br>(0.31) | 2.87<br>(0.33) | 2.91<br>(0.28) | 2.64<br>(0.34) |
| Inferior Temporal<br>Cortex Thickness | 2.68<br>(0.12) | 2.60<br>(0.22) | 2.57 (0.17) | 2.54 (0.15) | 2.57<br>(0.19) | 2.57<br>(0.23) | 2.57<br>(0.15) | 2.53<br>(0.09) |
| Parietal Cortex<br>Thickness | 2.25<br>(0.09) | 2.15<br>(0.09)<sup>a</sup> | 2.25 (0.11)<sup>b</sup> | 2.11 (0.14) | 2.26<br>(0.13) | 2.02<br>(0.18) | 2.25<br>(0.08) | 2.16<br>(0.08) |

### Regional differences in [¹⁸F]RO948 binding across sporadic Alzheimer’s disease continuum

Figure 1 shows individual tau-PET scans in the 9 MCI-EOAD and 7 MCI-LOAD patients, illustrating marked variability in binding across the MCI Aβ+ group. Individuals were categorized according to the regional extent of [¹⁸F]RO948 binding, reflecting progressive stages of tau accumulation from medial temporal to neocortical regions. Among MCI-EOAD individuals, five showed early binding in the medial-temporal regions, one displayed lateral temporal intermediate binding, and three exhibited advanced neocortical involvement. In contrast, MCI-LOAD patients showed three cases with medial temporal early binding and four with lateral temporal intermediate tau-PET binding. Overall, MCI-EOAD cases exhibited a greater spatial spread and burden of tau than MCI-LOAD (Fig. 1); this was also evident in EOAD vs LOAD individuals (fig. S1).

**Fig. 1.**
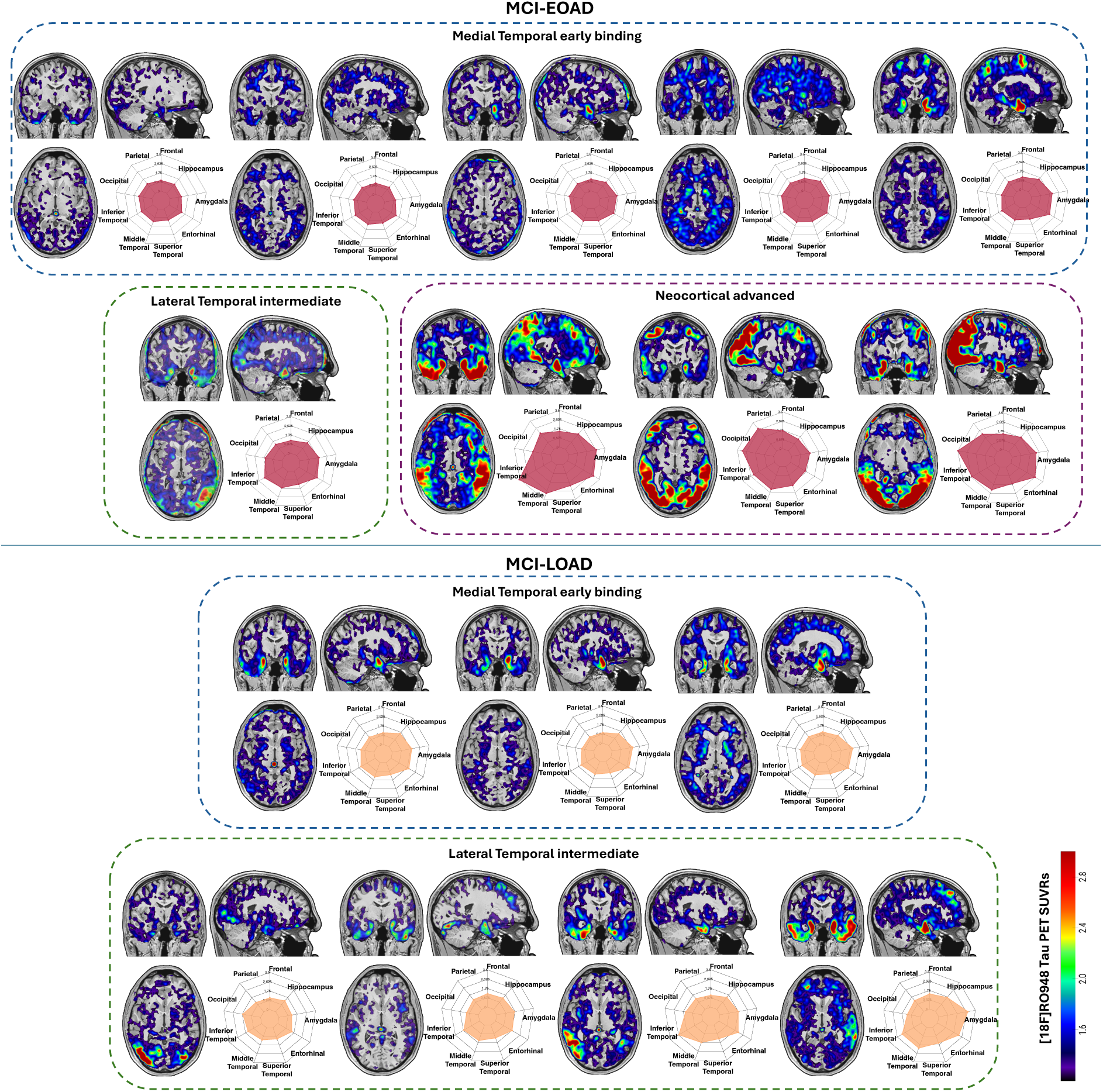
[18F]RO948 tau binding distribution in MCI groups by age at onset. [¹⁸F]RO948 tau PET scans (overlayed on corresponding MRI for anatomical reference) for Mild Cognitive Impairment amyloid-positive (MCI Aβ+) individuals are shown and subdivided according to early-onset (MCI-EOAD) and late-onset (MCI-LOAD) Alzheimer’s disease subgroups. For each subject, coronal, sagittal and axial views and radar plots display Standard Uptake Value Ratio (SUVR) values across key brain regions (Frontal, Parietal, Occipital, Inferior Temporal, Middle Temporal, Superior Temporal, Entorhinal, Amygdala, Hippocampus). Individuals were categorized according to the regional extent of [¹⁸F]RO948 binding, reflecting progressive stages of tau accumulation from medial temporal to neocortical regions.

Figure 2 shows the regional differences in [¹⁸F]RO948 tau-PET Standardized Uptake Value Ratio (SUVRs) across AD diagnostic groups. MCI Aβ+ individuals exhibited significantly higher tau-PET SUVRs in medial-temporal regions, including the entorhinal cortex, amygdala, and hippocampus (all *p* < 0.001), and extended also to inferior temporal region (*p* < 0.01) relative to cognitively normal (CN) participants. No significant differences in these regions were observed between MCI-EOAD and MCI-LOAD. When involving neocortical regions, overall AD individuals showed higher tau-PET levels than CN and MCI Aβ+ (Fig. 2), but group-specific differences emerged: the EOAD group (both MCI and AD), demonstrated a wider range of tau-PET SUVRs compared with the LOAD group. Specifically, parietal binding was significantly higher in EOAD than LOAD (*p* =0.046) (Fig. 3 and fig. S2). Voxel-wise analyses (Fig. 3) confirmed these regional findings, showing increased binding in medial-temporal regions in both MCI-EOAD and MCI-LOAD relative to CN. In EOAD and LOAD patients, the tau-PET binding extended to neocortical areas but with a distinct regional involvement. No differences were found in tau-PET SUVR between MCI Aβ- and CN groups. Region-specific tau-PET SUVRs are provided in Table 1 and Table S1.

**Fig. 2.**
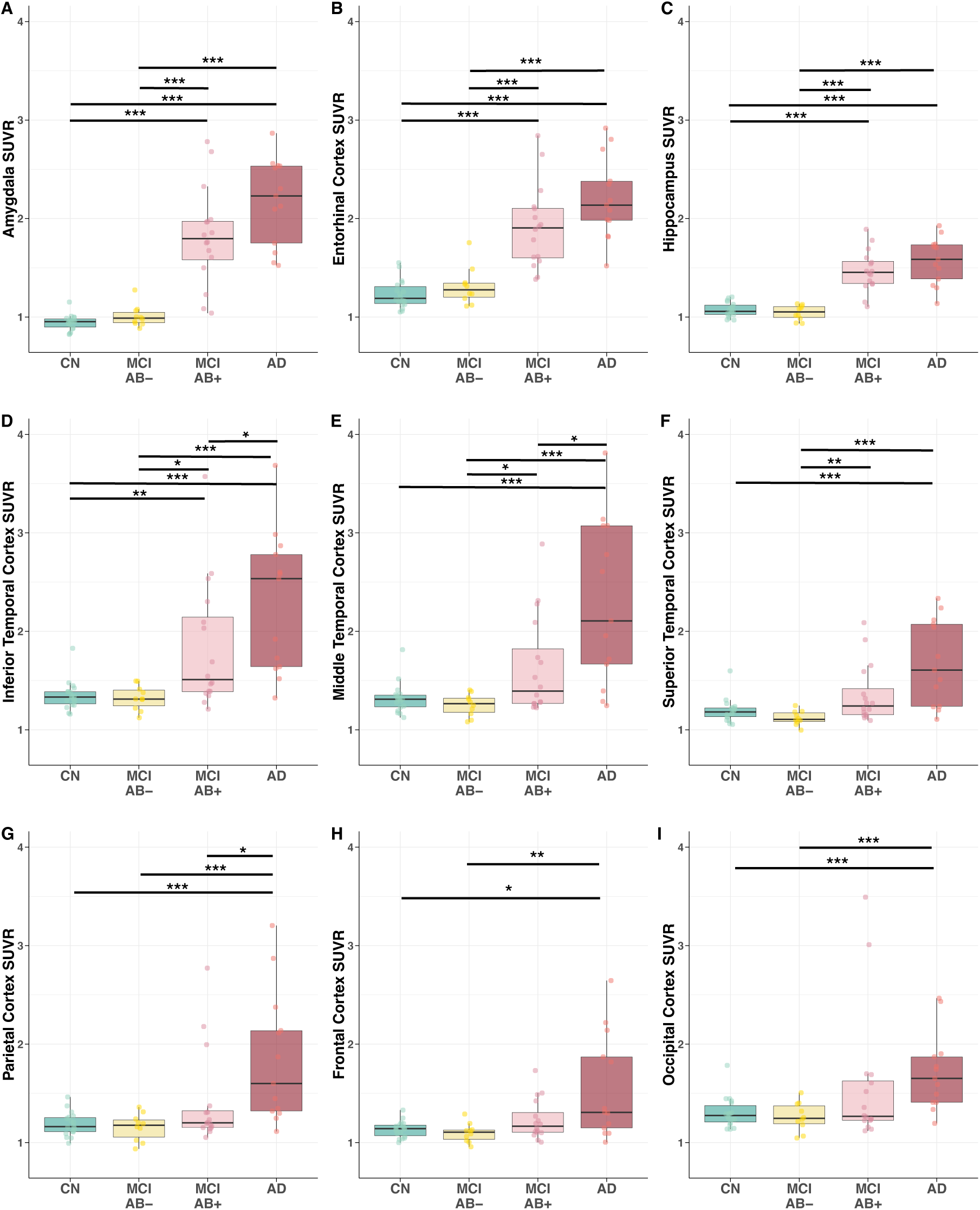
Regional differences in [¹⁸F]RO948 tau PET Standardized Uptake Value Ratio (SUVRs) across Alzheimer’s disease diagnostic groups. Boxplots showing Standardized Uptake Value Ratio (SUVR) values in the selected regions across cognitive normal (CN), mild cognitive impairment amyloid-negative (MCI Aβ-), mild cognitive impairment amyloid-positive (MCI Aβ+) and Alzheimer’s Disease (AD) groups.

**Fig. 3.**
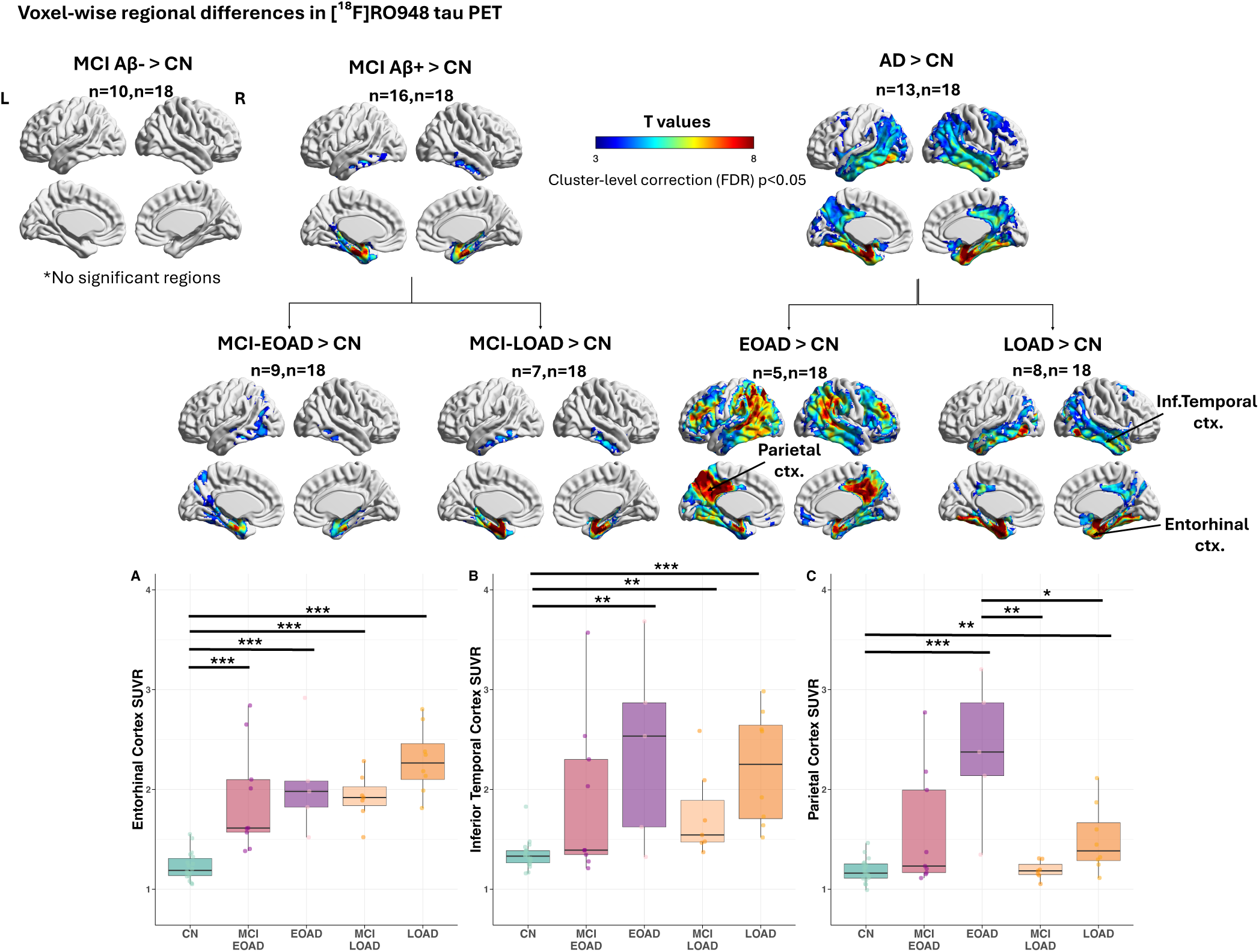
Voxel-wise and regional-specific differences in [¹⁸F]RO948 tau PET Standardized Uptake Value Ratio (SUVR) across Alzheimer’s disease diagnostic groups. Statistical parametric maps showing voxel-wise *t*-tests of [¹⁸F]RO948 tau-PET Standardized Uptake Value Ratio (SUVR) between each clinical group and the Cognitively Normal (CN) reference group. The voxel-wise t-tests were corrected at a cluster level with the FDR method at p < 0.05. Boxplots showing Standardized Uptake Value Ratio (SUVR) values in the selected regions across cognitively normal (CN), Mild Cognitive Impairment amyloid-positive (MCI Aβ+) and Alzheimer’s Disease (AD) groups in both early-onset (EOAD) and late-onset (LOAD) Alzheimer’s disease. Asterisks indicate significance levels from pairwise comparisons (\**p* < 0.05; \*\**p* < 0.01; \*\*\**p* < 0.001).

When we defined regional abnormal tau signal as z-scores>2 relatively to CN individuals the MCI Aβ+ and AD individuals showed highest percentage of abnormal [^18^F]RO948 tau-PET SUVR in the predominantly in the medial-temporal and limbic areas (i.e. the entorhinal cortex, the amygdala and the hippocampus) (fig. S3A). Overall, EOAD demonstrated -more frequently-abnormally high tau-PET binding in several brain areas compared with LOAD group (fig. S3B-C).

### Group differences in fluid biomarker levels across Alzheimer’s disease subtypes

Significant increases in p-tau217, p-tau181, p-tau231, p-tau217/Aβ42, GFAP and NFL levels were observed in both MCI Aβ+ and AD patients compared with CN individuals (all *p’s* < 0.05) (fig. S4). Figure 4 shows how plasma p-tau217 and GFAP levels were higher in both EOAD and LOAD individuals (MCI Aβ+ and AD) compared with CN (all *p* < 0.05). Plasma NFL showed only increased levels in LOAD (MCI Aβ+ and AD) compared with CN (all *p* < 0.01). No significant differences were observed between EOAD and LOAD within either the MCI Aβ+ or AD groups. Plasma Aβ42/40 levels did not differ significantly between groups (Fig. 4, Table 1). Plasma levels’ means and SD are depicted in Table 1 and Table S1.

**Fig. 4.**
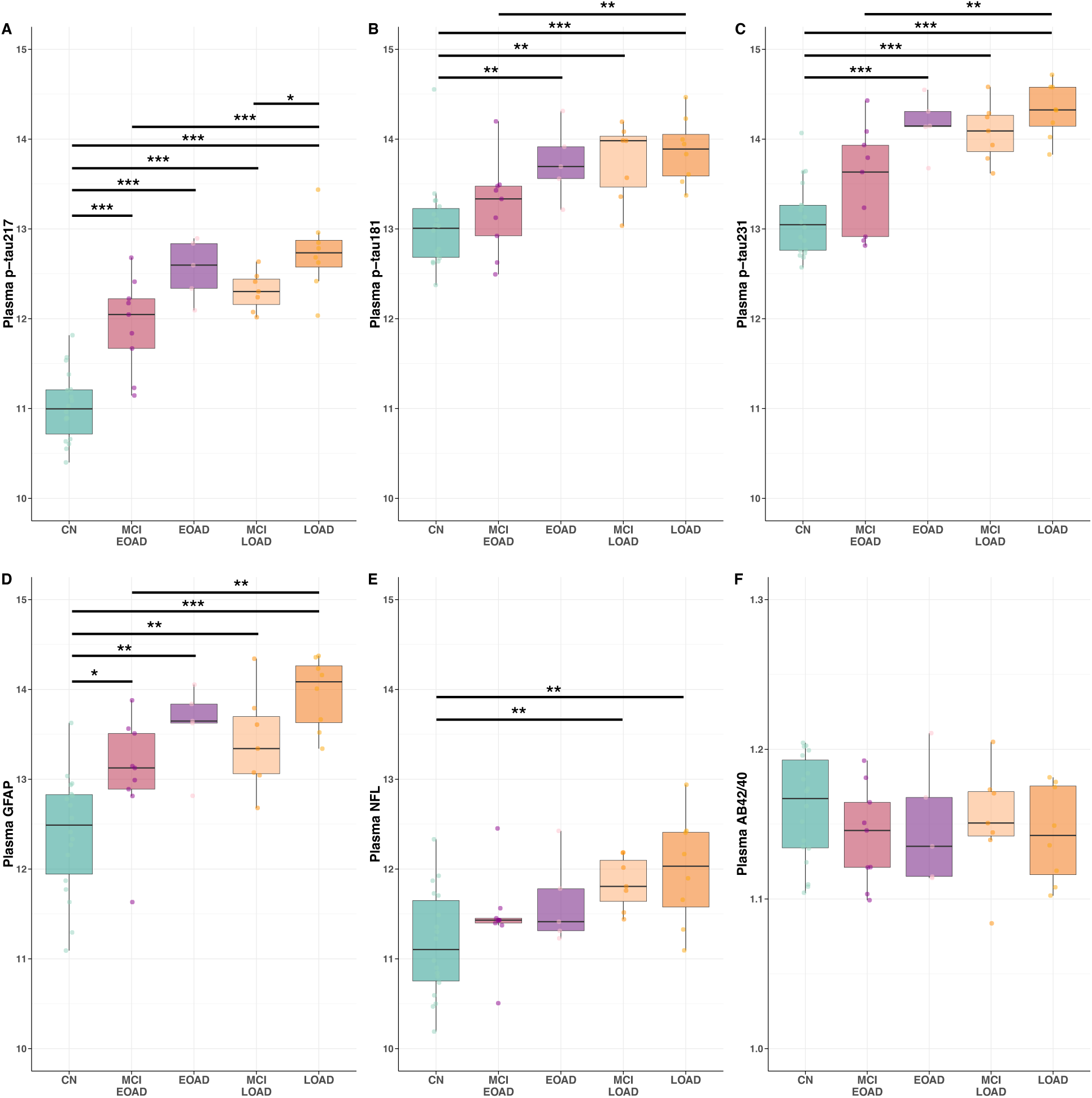
Plasma biomarker differences across Alzheimer’s disease diagnostic groups. Boxplots show the distribution of six plasma biomarkers across Alzheimer’s Disease diagnostic groups: cognitively normal (CN), early-onset and late-onset with Mild Cognitive Impairment (MCI Aβ+) or Alzheimer’s Disease (AD). Panels display plasma levels of phosphorylated tau:**(A)** p-tau217, **(B)** p-tau181, **(C)** p-tau231 and **(D)** Glial fibrillary acidic protein (GFAP), **(E)** neurofilament light chain (NFL), and **(F)** the plasma amyloid-beta (Aβ42/40) ratio. Individual data points are shown with overlaid boxplots representing the median and interquartile range (IQR); whiskers extend to 1.5×IQR. Horizontal lines indicate significant pairwise comparisons between groups based on post hoc tests, adjusted for multiple comparisons. Significance levels are denoted as follows: (\**p* < 0.05; \*\**p* < 0.01; \*\*\**p* < 0.001). NULISA biomarkers are expressed in NULISA Protein Quantification (NPQ) units.

### Age effects on tau and fluid biomarkers

When analyzing age as a continuous variable (fig. S5), a significant negative correlation was observed between the parietal [¹⁸F]RO948 SUVR and age within the AD and MCI Aβ+ groups (*rho* = –0.47, *p* = 0.050), while adjusting for cognitive status (MMSE-adjusted *partial rho* = –0.51, *p* = 0.045). No such correlation was present in CN individuals. In contrast, plasma biomarkers significantly increased with age across the total sample, particularly p-tau217, p-tau181, p-tau231, GFAP, and NFL (*partial rho* 0.68 to 0.36, *p* < 0.05), independent of cognitive status (MMSE-adjusted).

### Cross-sectional associations between tau-PET and fluid biomarkers

Plasma p-tau217 was the plasma biomarker most strongly correlated with tau-PET SUVRs, with stronger positive correlations in medial-temporal regions (i.e., the amygdala, the entorhinal cortex, and the hippocampus), followed by the temporal and parietal cortices (Fig. 5, fig. S6). GAM curves indicated increasingly non-linear associations in the inferior temporal and the parietal regions compared with the entorhinal cortex, as also evidenced in fig. S7. Plasma p-tau181 showed positive correlations with medial-temporal regions, whereas plasma p-tau231 extended also to correlate with lateral temporal regions (fig. S6).

**Fig. 5.**
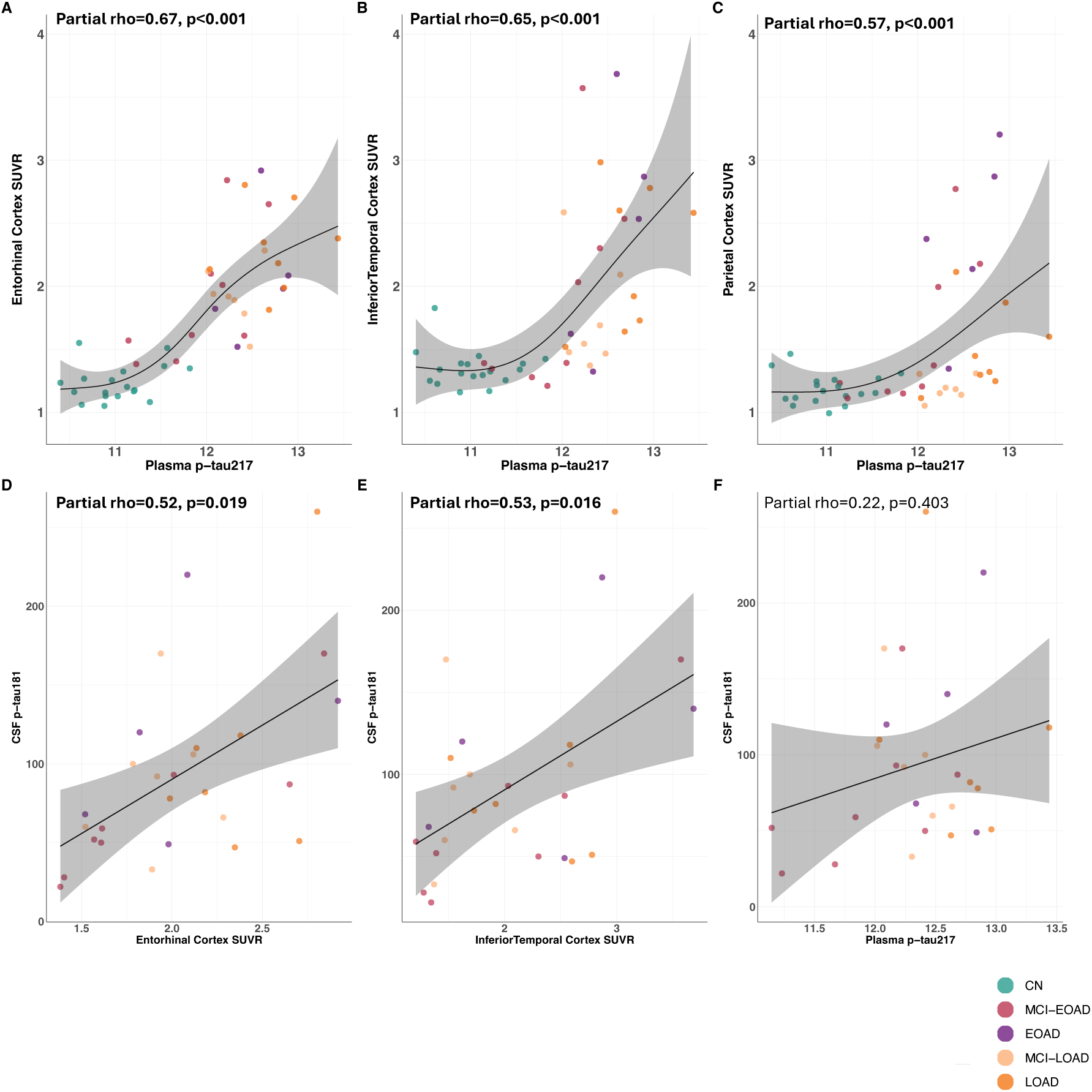
Regional correlations between [¹⁸F]RO948 tau-PET binding and plasma p-tau217 and their correlation with CSF p-tau181 across Alzheimer’s disease diagnostic groups. Panels **(A-C)** display associations between tau-PET Standard Uptake Value Ratio (SUVR) and plasma phosphorylated tau (p-tau217) in main regions explored of **(A)** entorhinal, **(B)** inferior temporal, **(C)** and parietal cortices. Panels **(D-E)** display correlations between Cerebrospinal Fluid (CSF) phosphorylated tau (p-tau181) and tau-PET SUVR, **(F)** correlations between CSF p-tau181 and plasma p-tau217. Smooth curves were fitted using generalized additive models (GAMs) with a Gamma distribution and log link **(A-C)** or linear models (LMs)**(D-F)**. Shaded areas indicate 95% confidence intervals. Colored dots indicate individual data points across five groups: cognitively normal (CN), early-onset and late-onset mild cognitive impairment (MCI-EOAD/MCI-LOAD) or Alzheimer’s Disease (EOAD/LOAD). Partial Spearman’s rho and FDR-corrected p-values are reported within each panel (n=47). Partial correlation coefficients (rho) and p-values are derived from models controlling for Sex and APOE status. NULISA plasma p-tau217 levels are expressed in NULISA Protein Quantification (NPQ) units.

Tau-PET showed a significant positive correlation with CSF p-tau181 in the entorhinal (*partial rho* = 0.52; *p* = 0.019) and the inferior temporal cortex (*partial rho* = 0.53; *p* = 0.016). Plasma p-tau217, as well as the other plasma biomarkers tested, did not show significant correlations with any CSF biomarker (Fig. 5 and fig. S6).

### Cross-sectional associations between tau-PET and fluid biomarkers by age at onset subtypes

When examining associations between tau-PET and plasma p-tau217, in both EOAD and LOAD, smoothed terms were significant across ROIs. Associations were linear in the medial-temporal regions for both groups, while inferior temporal and parietal regions showed linear patterns in LOAD and non-linear patterns in EOAD. A Wald test confirmed significant curve differences in the parietal cortex, driven by a strong non-linear association between markers in EOAD (fig. S7A–C).

No differences were observed between EOAD and LOAD for the association between tau-PET and CSF p-tau181. In contrast, the relationship between plasma p-tau217 and CSF p-tau181 showed a significant age group (EOAD vs LOAD) interaction; however, associations within each group were not significant (fig. S7D-I).

### Cross-sectional associations between tau-PET and fluid biomarkers with cognition

Significant negative correlations were observed between tau-PET binding and all cognitive domains assessed, except for attention. All results reported in the text below are shown in fig. S6.

The inferior temporal was the region most consistently correlated with cognitive performance across domains, including verbal and visuospatial episodic memory, executive function, and MMSE (all *p* < 0.05). The parietal region showed a strong negative correlation with visuospatial memory (*p* = 0.001) (Fig. 6 A-D). In contrast, plasma p-tau217 displayed weaker yet consistent negative correlations with MMSE, verbal and visuospatial episodic memory, but not executive function (all *p* < 0.05) (Fig. 6 E-H). Attention did not reveal any significant correlation either with tau-PET or plasma biomarkers.

**Fig. 6.**
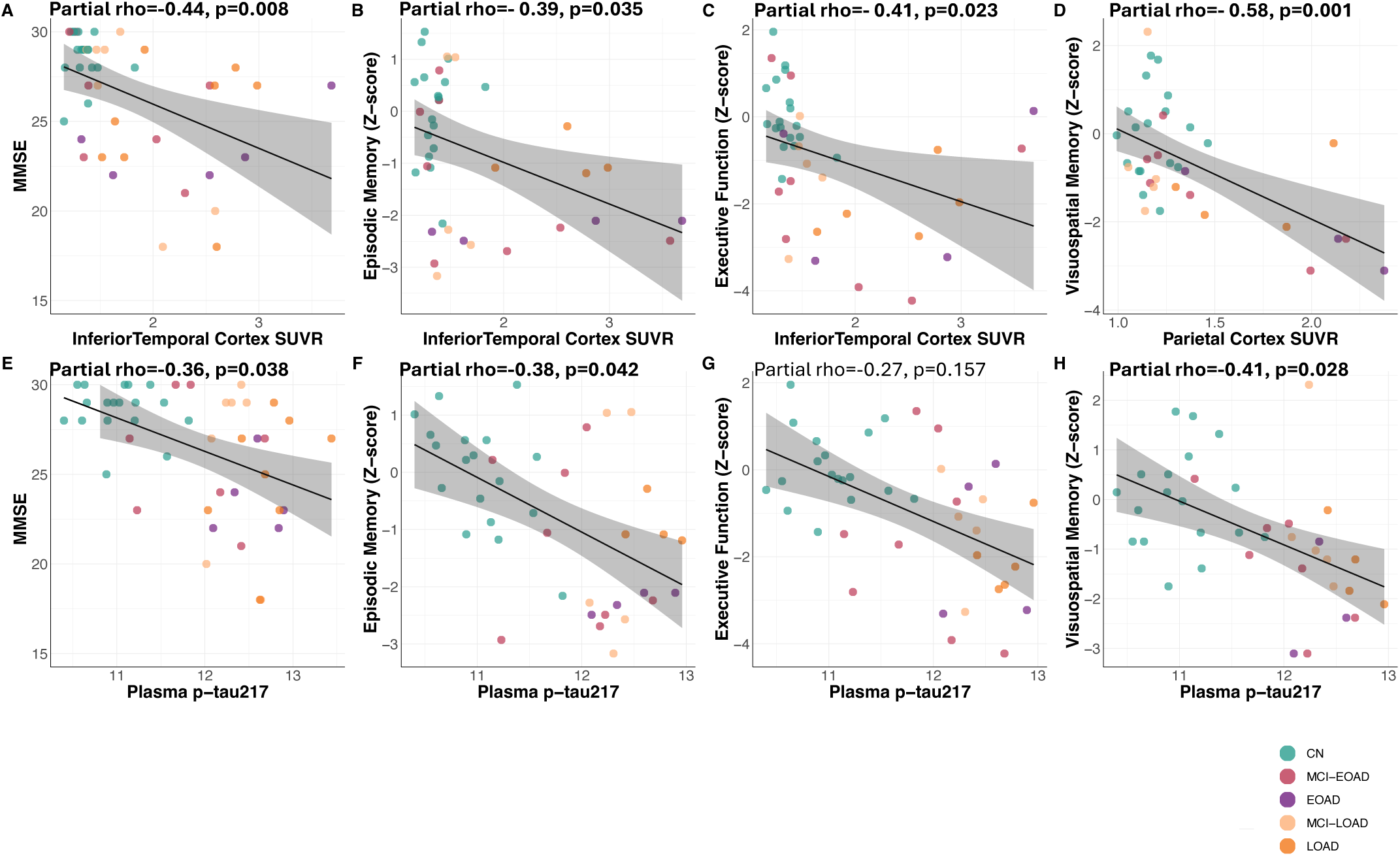
Regional correlations between [¹⁸F]RO948 tau PET binding, plasma biomarkers and cognition. (Panels A-D) Correlations between Inferior Temporal [¹⁸F]RO948 Standard Uptake Value Rario (SUVR) and **(A)** Mini-mental State Examination (MMSE), **(B)** Episodic Memory, **(C)** Executive Function, **(D)** and Parietal [¹⁸F]RO948 SUVR and Visuospatial Memory. **(Panels E-H)** Correlations between plasma phosphorylated tau (p-tau217) and **(E)** MMSE, **(F)** Episodic Memory, **(G)** Executive function, and **(H)** Visuospatial memory. MMSE(n=45), Episodic Memory z-score(n=39) Visuospatial Memory Z-score (n=36), Executive Function Z-score (n=40), and Attention Z-score(n=32). Shaded areas indicate 95% confidence intervals. Colored dots indicate individual data points across five groups: cognitively normal (CN), early-onset and late-onset with Mild Cognitive Impairment (MCI-EOAD/MCI-LOAD) or Alzheimer’s Disease (EOAD/LOAD). Partial Spearman’s rho and FDR-corrected p-values are reported within each panel derived from models controlling for Sex and APOE status. NULISA plasma p-tau217 levels are expressed in NULISA Protein Quantification (NPQ) units.

### Cross-sectional associations between tau-PET tau and plasma biomarkers with cognition by age at onset subtypes

Significant negative associations with visuospatial memory were observed in EOAD but not in LOAD, for both the amygdala (*β* = –1.78, *p* = 0.013) and the hippocampus (*β* = – 3.74, *p* = 0.012). No other significant interactions were observed (fig. S8).

When examining representative individuals across clinical groups, substantial heterogeneity in tau burden was evident within both EOAD and LOAD groups, which was reflected in divergent cognitive profiles. Notably, EOAD individuals tended to show poorer visuospatial memory performance relative to LOAD participants, aligning with the distribution of tau pathology (fig. S9).

### Cross-sectional associations between tau-PET tau and plasma biomarkers with brain atrophy by age at onset subtypes

Similar negative correlation were observed between cortical thickness or volume measures and tau-PET or plasma biomarkers. Both tau-PET measures and plasma p-tau217 biomarkers showed significant negative correlations with medial-temporal and lateral temporal MRI measures (*all p* < 0.05). Lower parietal thickness was negatively associated with higher parietal tau-PET SUVRs (*β* = -0.202, *p* = 0.001) and higher plasma p-tau217 (*β* = -0.107, *p* = 0.035) in the EOAD group specifically (Fig. 7). No other significant differences between EOAD vs LOAD were found in the tau-PET/plasma-thickness relationships (fig. S10).

**Fig. 7.**
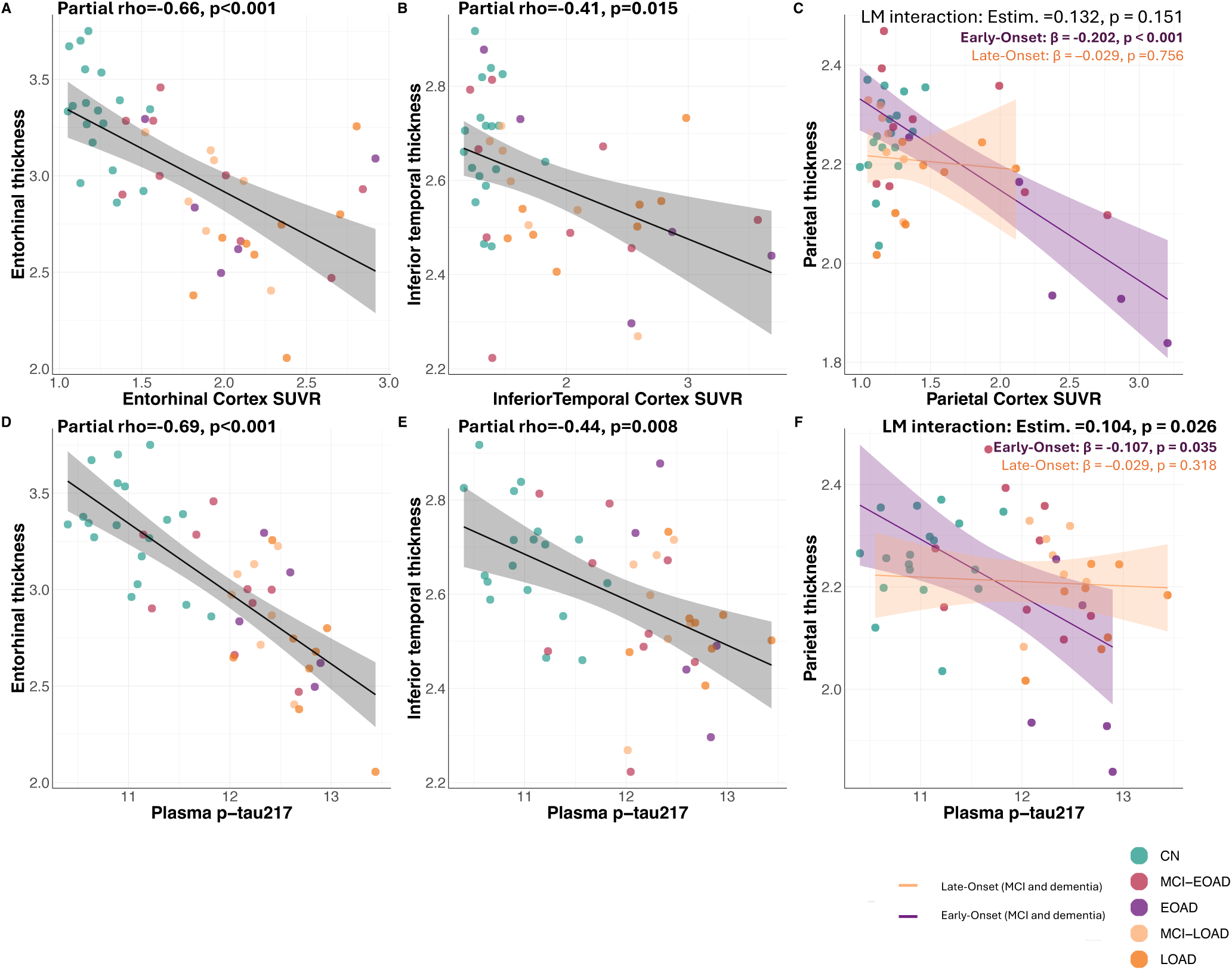
Regional correlations between [¹⁸F]RO948 tau PET binding, plasma biomarkers and cortical thickness. Panels (C).Correlations between: **(A)** Entorhinal [¹⁸F]RO948 SUVR and Entorhinal thickness, **(B)** Inferior Temporal [¹⁸F]RO948 SUVR and Inferior Temporal thickness, **(C)** Parietal [¹⁸F]RO948 SUVR and Parietal thickness, **(Panels D-F)** Correlations between plasma phosphorylated tau (p-tau217) and: **(D)** Entorhinal thickness, **(E)** Inferior Temporal thickness, **(F)** Parietal thickness. Scatter plots show raw data points and unadjusted regression lines. Shaded areas represent 95% confidence intervals. Partial correlation coefficients (rho) and p-values are derived from models controlling for Sex and APOE status. Panel **(C, F)** fitted lines are presented by EOAD/LOAD and LM interaction estimates and p-values are derived from models controlling for Sex and APOE status. Group abbreviations: CN = cognitively normal; MCI-EOAD/MCI-LOAD= early-onset and late-onset disease-Mild Cognitive Impairment; EOAD/LOAD = early-onset and late-onset Alzheimer’s disease.

## DISCUSSION

In this study, we characterized the regional distribution of tau pathology using the second-generation tau-PET tracer [¹⁸F]RO948 in a clinically representative cohort of sporadic Alzheimer’s disease patients compared to cognitive healthy controls across the disease continuum. We further examined how tau-PET relates to plasma biomarkers, cognitive performance and brain atrophy, with a particular focus on disease stage and age at onset (comparing EOAD vs LOAD). Although differences in tangle burden have been linked to both disease stage and age at onset (EOAD/LOAD), it remains unclear how such variation is reflected in tau-PET measures and fluid biomarkers (plasma or CSF tau levels). Regional tau accumulation first appeared in medial-temporal regions and was already elevated in MCI Aβ+. MCI-EOAD individuals showed greater neocortical involvement, and EOAD (AD) patients displayed higher parietal tau-PET binding than LOAD (AD), suggesting that early-onset forms exhibit more widespread cortical deposition even at prodromal stages. While plasma p-tau217 levels varied with disease stage, they did not differ by age at onset. In EOAD, the elevated tau burden measured by PET showed a non-linear association with plasma p-tau217 levels, which plateaued even as tau-PET binding continued to increase. In contrast, LOAD showed more gradual, linear associations. Both tau-PET and plasma p-tau217 were negatively related to cognition and cortical thickness, although the associations were stronger and region-specific for tau-PET.

We observed early tau accumulation in MCI Aβ+ individuals compared to cognitively normal participants, indicating that tau pathology associated with neurofibrillary tangles can be detected at earlier clinical stages than previously visualized by imaging. Specifically, subcortical structures such as the amygdala and hippocampus mirrored tau accumulation in the entorhinal cortex, consistent with postmortem data identifying these regions as the earliest sites of tangle formation(*5, 6, 25, 26*) These results support [¹⁸F]RO948 as a sensitive tracer capable of detecting initial stages of tau aggregation in the brain, including regions such as the hippocampus, where first-generation tracers like [¹⁸F]AV-1451 (Flourtaucipir) have shown limited signal-to-noise ratio.(*19, 27–29*) The improved sensitivity of [¹⁸F]RO948 and other second-generation tracers (e.g., [¹⁸F]MK6240) enhances discrimination at the prodromal stage of AD, where tau accumulation is subtle. In contrast, [¹⁸F]Flortaucipir shows substantially lower discriminative accuracy at this stage due to less pronounced tau binding in MCI than in AD dementia.(*30, 31*) Beyond the canonical Braak trajectory,(*5, 32, 33*) we identified a distinct topographical influence of age at onset: while both MCI-EOAD and MCI-LOAD showed elevated tau in medial temporal regions, EOAD cases exhibited greater neocortical involvement even at the MCI stage. These findings align with prior reports of regional heterogeneity by age at onset,(*8, 9, 34–36*) and underscore the value of second-generation tau-PET tracers like [¹⁸F]RO948 for mapping spatially variable pathology in vivo.

Recent efforts have explored whether plasma biomarkers, particularly p-tau217, can serve as surrogates for tau-PET in clinical settings for early detection(*37*) , patient stratification(*22*) and disease monitoring. Although initial results are promising, current clinical guidelines recommend caution, limiting blood-based biomarker use to patients with objective cognitive impairment assessed in specialized memory clinics.(*38*) Consistent with this framework we found that plasma p-tau217 levels increased progressively across diagnostic stages, differentiating CN, MCI Aβ+ and AD, highlighting its sensitivity to disease stage.(*39, 40*) Emerging evidence indicates that most of the variance in plasma p-tau217 levels is explained by amyloid-β pathology, whereas the remaining variance may reflect individual differences in tau accumulation(*41, 42*). Previous studies with [^18^F]Flortaucipir, demonstrated that plasma p-tau217 strongly correlates with temporo-parietal tau and outperforms other plasma biomarkers in predicting PET(*43, 44*) or autopsy tau positivity.(*45*) Recent studies using [¹⁸F]RO948 and [^18^F]MK6240 further confirm plasma p-tau217 as an early biomarker linked to early tau pathology. Specifically, in cognitively unimpaired individuals, plasma p-tau217 showed stronger associations with amyloid PET than with tau-PET positivity in the temporal meta-ROI. In contrast, in cognitively impaired individuals, plasma p-tau217 also identified tau-PET positivity.(*46, 47*) Importantly, elevated plasma p-tau217 abnormalities preceded PET-detectable tau, predicting early medial temporal tau accumulation.(*37*) These findings support the hypothesis that plasma p-tau217 rises early in an Aβ-dependent manner, preceding neocortical insoluble tau aggregation.(*37, 48*)

Extending these findings, we found that plasma p-tau217 did not differ significantly between MCI-EOAD and MCI-LOAD, nor between EOAD and LOAD groups. This contrasts with our tau-PET findings, which showed qualitatively higher SUVRs in MCI-EOAD. This apparent dissociation aligns with the regional correlation analyses: our results showed that plasma p-tau217 correlated most strongly with tau-PET in early-affected regions (the entorhinal cortex, hippocampus and amygdala). In later-affected neocortical regions—particularly the inferior temporal and parietal cortices—the relationship becomes increasingly non-linear, especially in EOAD, indicating divergence between soluble and aggregated tau measures. This pattern likely reflects a plateau of plasma p-tau217 levels once cortical tau aggregation accelerates, consistent with prior work showing that plasma p-tau217 rises early but saturates even as PET signal continues to increase.(*49*) These results suggest that EOAD may represent a subgroup not fully captured by plasma biomarkers. This finding has important implications for clinical trial design and participant stratification, as relying solely on plasma biomarkers could underestimate cortical tau burden in early-onset presentations.^65^ Our findings suggest that plasma p-tau217 capture early, amyloid-driven phosphorylation dynamics, but it does not reflect the full spatial spread of tau pathology measured by tau-PET —highlighting the continued value of tau-PET for accurate disease stratification and for assessing therapeutic efficacy in individuals with distinct regional tau burdens.(*50*)

CSF measures provided complementary information: both CSF p-tau and t-tau positively correlated with tau-PET but not with plasma measures.^67^ This supports the notion that CSF tau indices more directly reflect central tangle formation and aggregated tau burden.(*51*) Our findings align with the A/T/N framework, which considers both tau-PET and CSF p-tau as valid indicators of tau pathology.(*52*) Although prior studies have shown that plasma p-tau217 can perform comparably to CSF p-tau217 in identifying Aβ and tau-PET positive individuals,(*47, 53*) our data suggest that both CSF and plasma tau biomarkers capture distinct biological windows. Plasma p-tau217 may better capture amyloid-driven phosphorylation changes, while CSF p-tau181 is more tightly associated with ongoing tangle formation visualized by PET.

In this study, we investigated cognitive symptoms across distinct domains and found that both tau-PET and plasma p-tau217 biomarkers correlated with cognitive performance and cortical thickness, yet tau-PET showed stronger, domain-specific associations. Previous longitudinal studies demonstrated that tau-PET more closely tracks cognitive decline than plasma biomarkers, (*54*) supporting its role as a marker of downstream neurodegeneration.(*11*) In our cohort of individuals with varying age at onset, plasma p-tau217 related broadly to global cognition (MMSE) and episodic memory (verbal, and visuospatial), reflecting its link to early-phase pathology.(*54–56*) Tau-PET, however, revealed distinct regional–cognitive associations, consistent with previous reports in amyloid-positive MCI and AD individuals.(*35, 57*) We extended these findings by showing that, particularly in EOAD, tau accumulation in the middle temporal and parietal cortices corresponded to visuospatial impairment—an EOAD-typical cognitive feature.(*58, 59*) These findings emphasize the complementary but distinct roles of plasma and imaging biomarkers: plasma measures are informative for early detection, while tau-PET offers spatial precision to map cognitive phenotypes and disease severity.

A major strength of this work is that it is the first to directly assess plasma–tau-PET–cognition relationships using [¹⁸F]RO948 in both early-onset and late-onset Alzheimer’s disease across the AD continuum. The inclusion of both early- and late-onset cases enhances clinical representativeness. Nonetheless, several limitations must be acknowledged. The modest sample size and inclusion of both MCI and AD stages limited statistical power for subgroup analyses, although this heterogeneity mirrors real-world patient diversity. Correlations between CSF, plasma, and tau-PET biomarkers should be interpreted cautiously due to batch variability in CSF measurements. The lack of a cognitively normal group in CSF analyses to anchor our relationships may also have influenced the detection of certain correlations. The cross-sectional design precludes causal inference; future longitudinal studies should evaluate how plasma and PET biomarkers differentially predict cognitive and structural decline over time. Finally, emerging evidence suggests sex differences in tau deposition, with women showing higher tau accumulation.(*60, 61*) While our exploratory analyses did not reveal sex as a significant moderator, the trend toward higher values in women warrants further study in larger, sex-balanced cohorts.

## Conclusions

While both plasma p-tau217 and tau-PET reflected disease stage, as indicated by cognitive and functional impairment, tau-PET additionally captured inter-individual heterogeneity in the spatial spread and burden of tau pathology, particularly in advanced stages where plasma p-tau217 levels tended to plateau and showed limited variability. Therefore, tau-PET appears better suited for tracking disease progression and evaluating treatment effects in clinical trials, especially in EOAD individuals who exhibit higher tau-PET levels even in prodromal stages. Together, these results underscore the importance of tau-PET as a critical biomarker for assessing disease severity and support its central role in clinical trials aimed at evaluating disease-modifying therapies. Moreover, the distinct patterns of tau accumulation observed in early-onset versus late-onset AD highlight the need to account for age-related heterogeneity when designing clinical trials and precision treatment strategies. Integrating plasma, CSF, and tau-PET measures—while considering age-at-onset and cognitive phenotype—will be essential for refining diagnostic algorithms and improving stratification in disease-modifying trials.

## Supplementary material

The PDF includes:

Figs. S1 to S10

Table S1

## Acknowledgements

We would like to express our gratitude to all participants in the PET studies and their families, as well as to the staff at the Cognitive Assessment Unit at Karolinska University Hospital Huddinge, the Uppsala PET Centre, Academical Hospital, Uppsala, and Clinical Neurochemistry Laboratory, Sahlgrenska University Hospital, Gothenburg. We thank F.Hoffmann-La Roche Ltd, Basel, Switzerland for providing the precursor for synthesis of [18F]RO948.

## Data availability

The data that support the findings of this study are available from the corresponding author upon reasonable request. Due to participant privacy and ethical restrictions, individual-level imaging and clinical data cannot be made publicly available.

