## Supplementary Figures and Table for "Regional [^18^F]RO948 tau-PET across the AD continuum in relation to plasma biomarkers, cognition and atrophy": Supplementary Figures and table_Preprint_v1.pdf

| Variable | CN<br>N = 18 | MCI Aß-<br>N = 10 | MCI Aß+<br>N = 16 | AD<br>N = 13 | MCI-EOAD<br>N = 9 | EOAD<br>N = 5 | MCI-LOAD<br>N = 7 | LOAD<br>N = 8 |
|---|---|---|---|---|---|---|---|---|
| Hippocampus SUVR | 1.07 (0.07) | 1.04 (0.08) | 1.47 (0.21) [a] | 1.57 (0.24) [a] | 1.47 (0.26) | 1.61 (0.23) | 1.46 (0.14) | 1.55 (0.25) |
| Amygdala SUVR | 0.94 (0.08) | 1.01 (0.11) | 1.82 (0.49) [a] | 2.17 (0.44) [a] | 1.76 (0.54) | 2.00 (0.45) | 1.89 (0.47) | 2.28 (0.42) |
| Frontal Cortex SUVR | 1.13 (0.08) | 1.10 (0.10) | 1.23 (0.20) | 1.55 (0.53) [a] | 1.28 (0.24) | 1.80 (0.63) | 1.17 (0.14) | 1.40 (0.42) |
| Occipital Cortex SUVR | 1.30 (0.16) | 1.26 (0.15) | 1.58 (0.69) | 1.71 (0.39) [ab] | 1.78 (0.86) | 1.95 (0.53) | 1.31 (0.21) | 1.57 (0.19) |
| Plasma p-tau231 | 13.10 (0.41) | 13.36 (0.52) | 13.77 (0.56) [a] | 14.26 (0.31) [ab] | 13.52 (0.59) | 14.16 (0.32) | 14.08 (0.33) | 14.32 (0.30) |
| Plasma p-tau181 | 13.02 (0.48) | 13.32 (0.72) | 13.46 (0.53) [a] | 13.82 (0.37) [a] | 13.23 (0.52) | 13.74 (0.41) | 13.74 (0.43) | 13.87 (0.37) |
| Plasma p-tau217/Aβ42 | 0.82 (0.04) | 0.85 (0.03) | 0.93 (0.04) [a] | 0.97 (0.02) [a] | 0.91 (0.03) | 0.97 (0.01) [c] | 0.94 (0.04) | 0.96 (0.03) |
| Plasma GFAP | 12.39 (0.66) | 13.41 (0.44) | 13.21 (0.61) [a] | 13.82 (0.45) [ab] | 13.06 (0.64) | 13.60 (0.47) | 13.41 (0.55) | 13.96 (0.40) |
| Plasma NFL | 11.16 (0.59) | 12.36 (0.76) | 11.62 (0.45) | 11.85 (0.58) [a] | 11.45 (0.49) | 11.63 (0.49) | 11.84 (0.30) | 11.99 (0.62) |
| Hippocampal Volume/TIV | 2.65 (0.33) | 2.08 (0.36) | 2.30 (0.37) [a] | 2.04 (0.31) [a] | 2.40 (0.38) | 2.06 (0.21) | 2.17 (0.34) | 2.03 (0.37) |
| Amygdala Volume/TIV | 1.02 (0.16) | 0.80 (0.15) | 0.84 (0.16) [a] | 0.72 (0.15) [a] | 0.89 (0.19) | 0.70 (0.13) | 0.78 (0.09) | 0.74 (0.17) |

**Table S1. Biomarker and atrophy data for the different study groups.** Values are presented as Mean (SD). Shown are SUVR values in the selected brain regions, plasma biomarker levels, and volumetric ratios across cognitively normal (CN), mild cognitive impairment amyloid-negative (MCI Aβ–), mild cognitive impairment amyloid-positive (MCI Aβ+), and Alzheimer's disease (AD) groups, for both early-onset (EOAD) and late-onset (LOAD) cases.Group comparisons displayed: CN vs MCI Aβ+ vs AD, MCI-EOAD vs MCI-LOAD, EOAD vs LOAD.

a = statistically different from CN, b = statistically different from MCI AB+.

c = statistically different from MCI-EOAD, d = statistically different from EOAD. Significance level $p<0.05$. **Abbreviations:** SUVR = Standardized Uptake Value Ratio; p-tau = phosphorylated tau;GFAP = glial fibrillary acidic protein;NFL = neurofilament light chain;TIV = total intracranial volume.

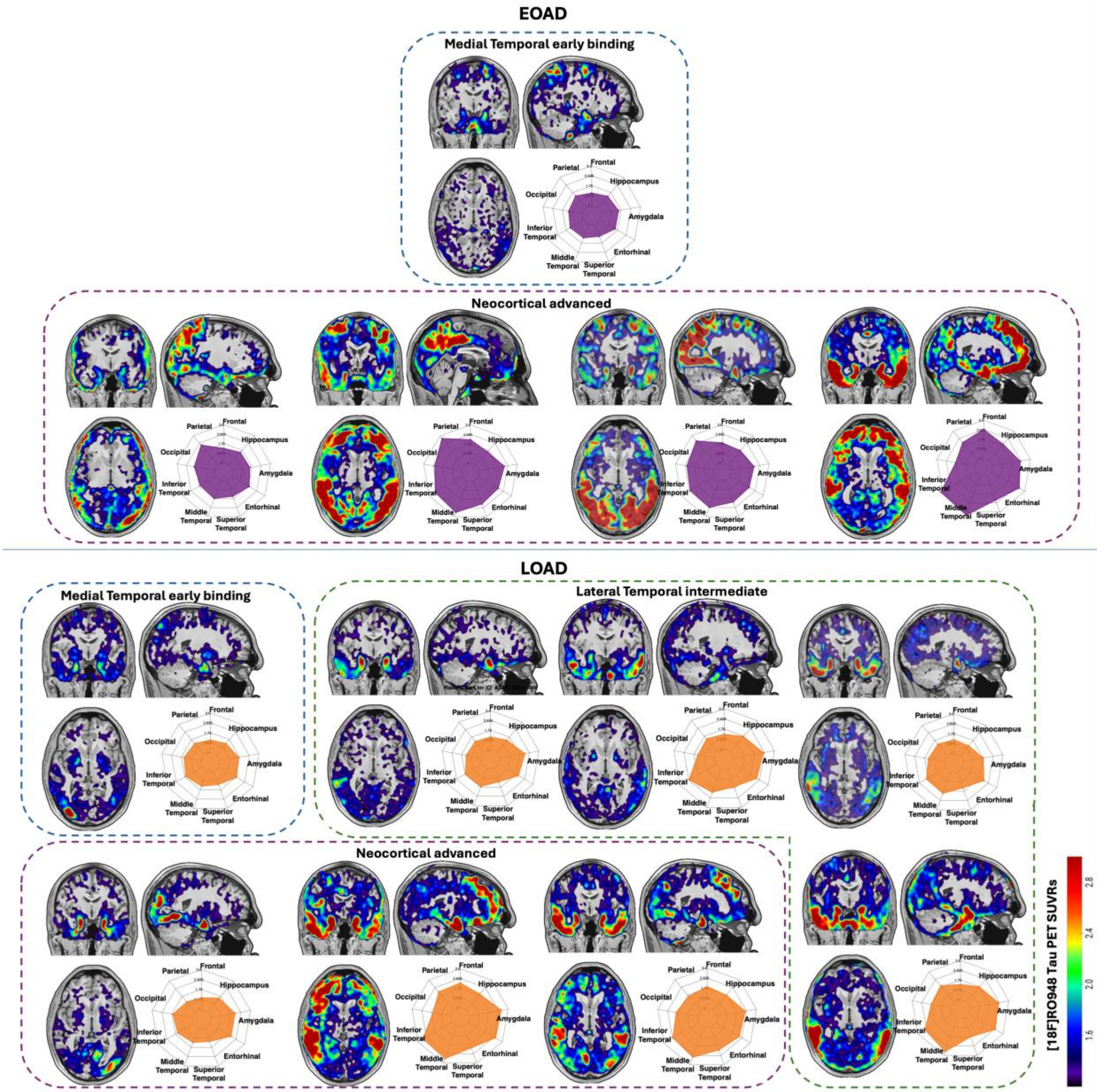


**Fig. S1. [18F]RO-948 TAU binding distribution in AD groups by age-at onset.** [$^{18}$F]RO948 tau PET scans (overlayed on corresponding MRI for anatomical reference) for Alzheimer's Disease (AD) individuals are shown and subdivided according to Early- (EOAD) and Late-Onset (LOAD) subgroups. For each subject, coronal, sagittal and axial views and radar plots display Standardized Uptake value Ratio (SUVR) values across key brain regions (Frontal, Parietal, Occipital, Inferior Temporal, Middle Temporal, Superior Temporal, Entorhinal, Amygdala, Hippocampus). Individuals were categorized according to the regional extent of [$^{18}$F]RO948 binding, reflecting progressive stages of tau accumulation from medial temporal to neocortical regions.

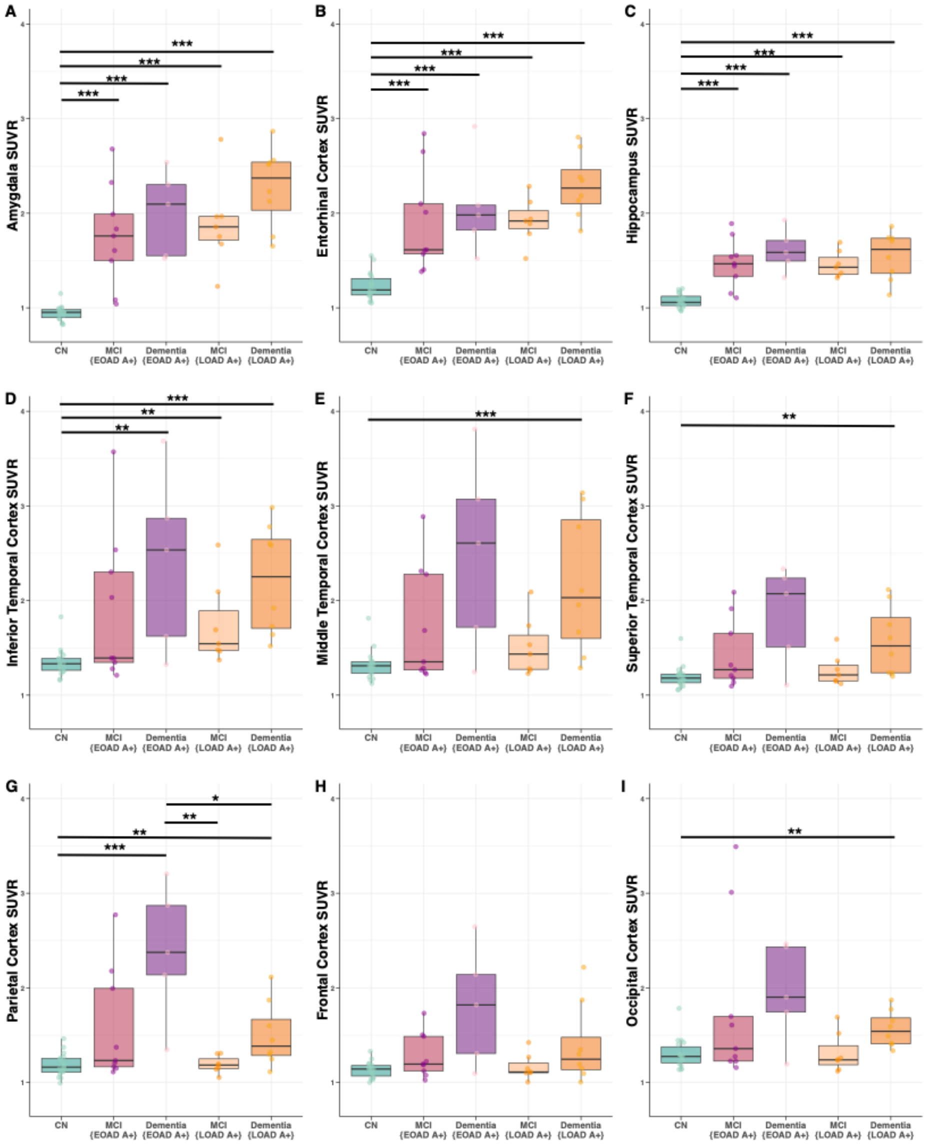


**Fig. S2. Regional differences in [18F]RO948 tau PET Standardized Uptake Value Ratios (SUVRs) across Alzheimer's disease diagnostic groups.** Boxplots showing Standardized Uptake Value Ratios (SUVRs) values in the selected regions across Cognitively Normal (CN) , Mild Cognitive Impairment (MCI) and Alzheimer's disease (AD) groups, for early-onset (EOAD) and late-onset (LOAD) subgroups. Asterisks indicate significance levels from pairwise comparisons (*$p < 0.05$; **$p < 0.01$; ***$p < 0.001$).

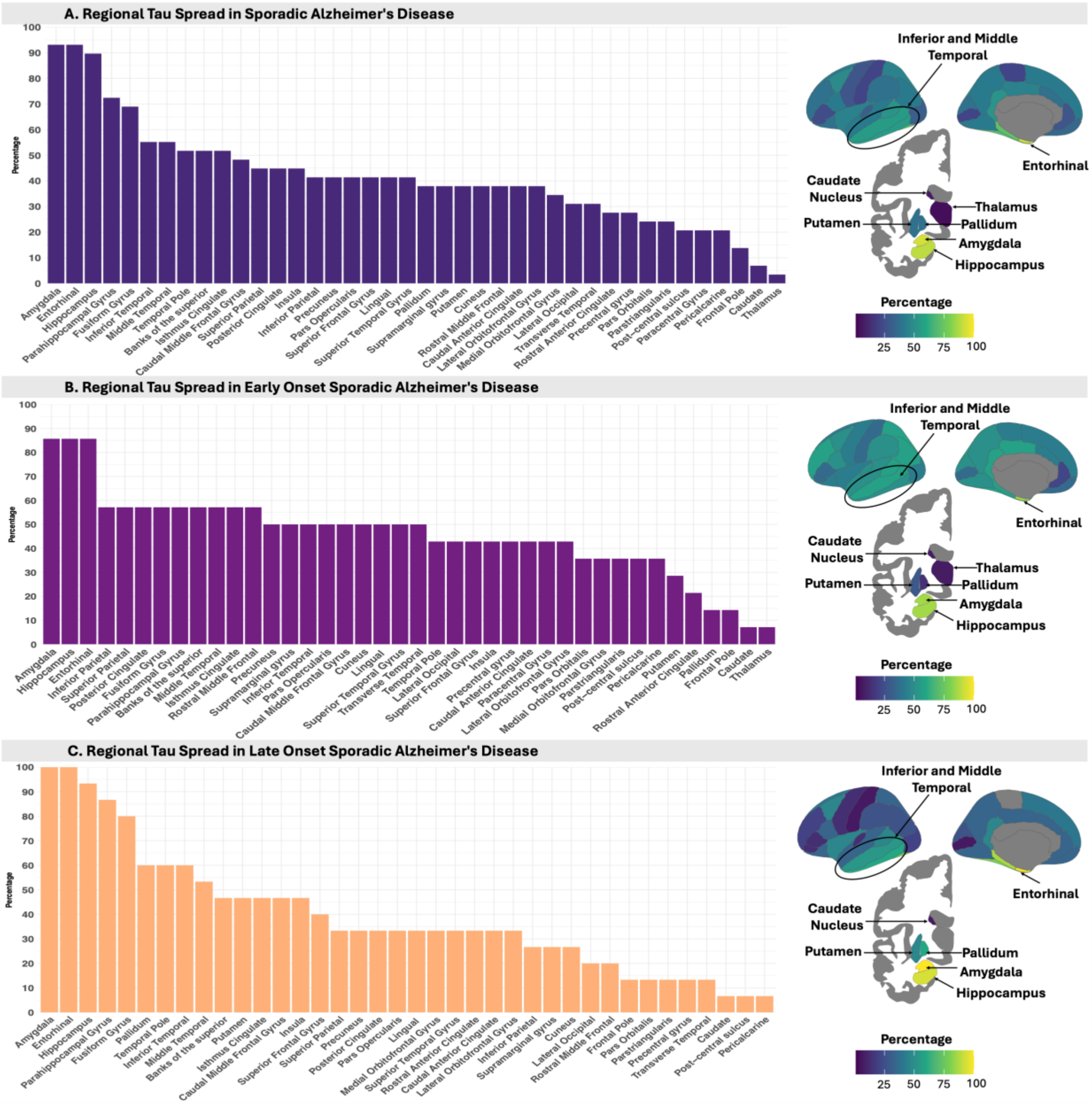


**Fig. S3. Regional Tau Spread in Sporadic Alzheimer's Disease.** The figure illustrates the percentage of individuals with abnormal [$^{18}$F]RO948 tau PET SUVRs across different brain regions, based on z-scores > 2 relative to cognitively normal (CN) controls. Top bar plots show the overall percentage of regional tau involvement among participants with Mild Cognitive Impairment (MCI) and Alzheimer's Disease (AD). The middle and bottom panels further divide the cohort into early-onset and late-onset subgroups. On the right, corresponding brain maps display the spatial distribution of regional tau involvement. Brighter colors indicate a higher proportion of individuals with tau-PET positivity in each region.

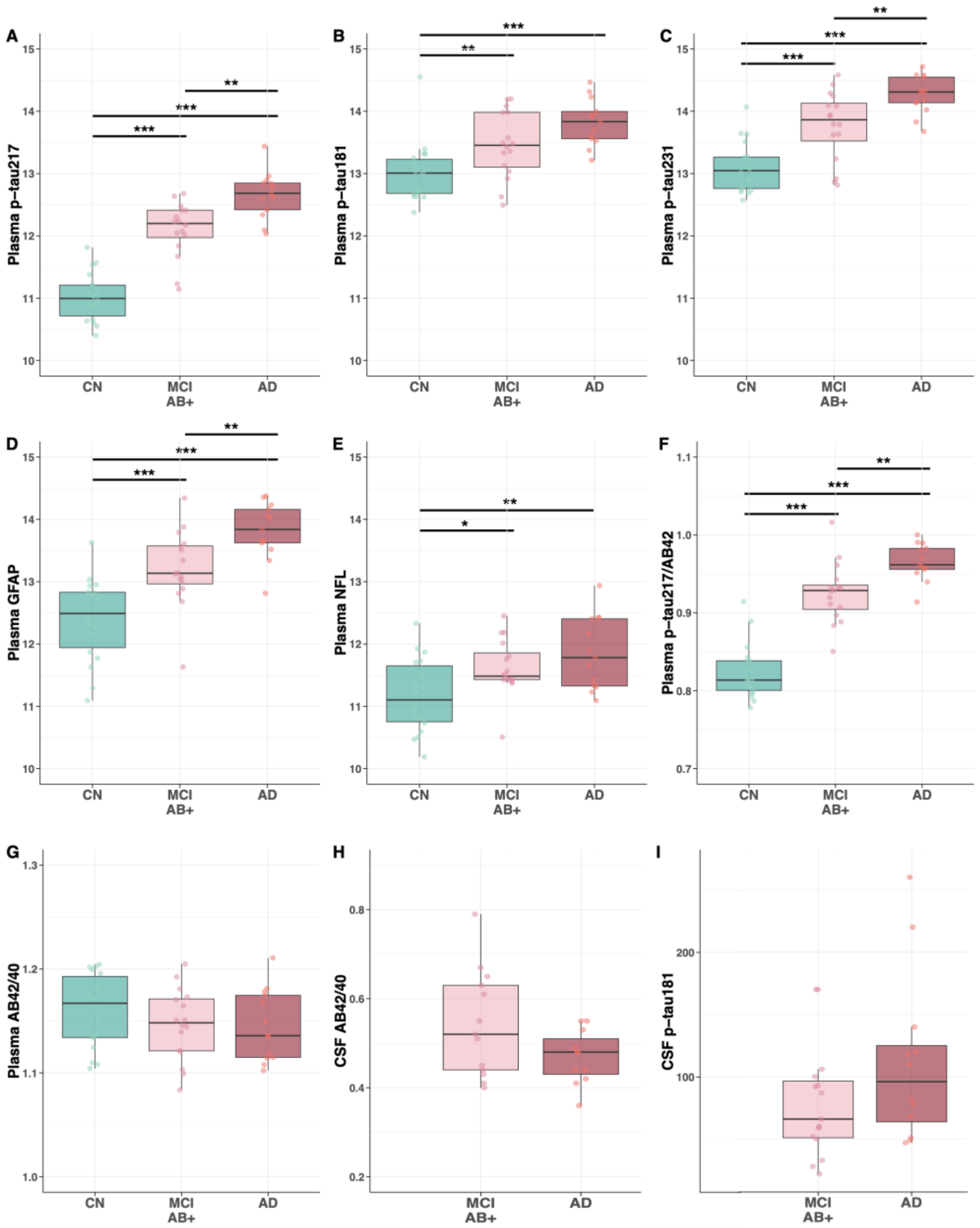


**Fig. S4. Plasma and CSF biomarker differences across Alzheimer's disease diagnostic groups.**Boxplots show the distribution of six plasma biomarkers across Alzheimer's Disease diagnostic groups: cognitively normal (CN), mild cognitive impairment amyloid-positive (MCI Aβ+), or Alzheimer's disease (AD). Panels display plasma levels of: phosphorylated tau**(A)** p-tau217, **(B)** p-tau181, **(C)** p-tau231, and **(D)** Glial fibrillary acidic protein (GFAP), **(E)** neurofilament light chain (NFL), and **(F)** the p-tau217/Aβ42 ratio. Individual data points are shown with overlaid boxplots representing the median and interquartile range (IQR); whiskers extend to 1.5×IQR. Horizontal lines indicate significant pairwise comparisons between groups based on post hoc tests, adjusted for multiple comparisons. Significance levels are denoted as follows: (*p < 0.05; ***p* < 0.01; ****p* < 0.001). NULISA biomarkers are expressed in NULISA Protein Quantification (NPQ) units.

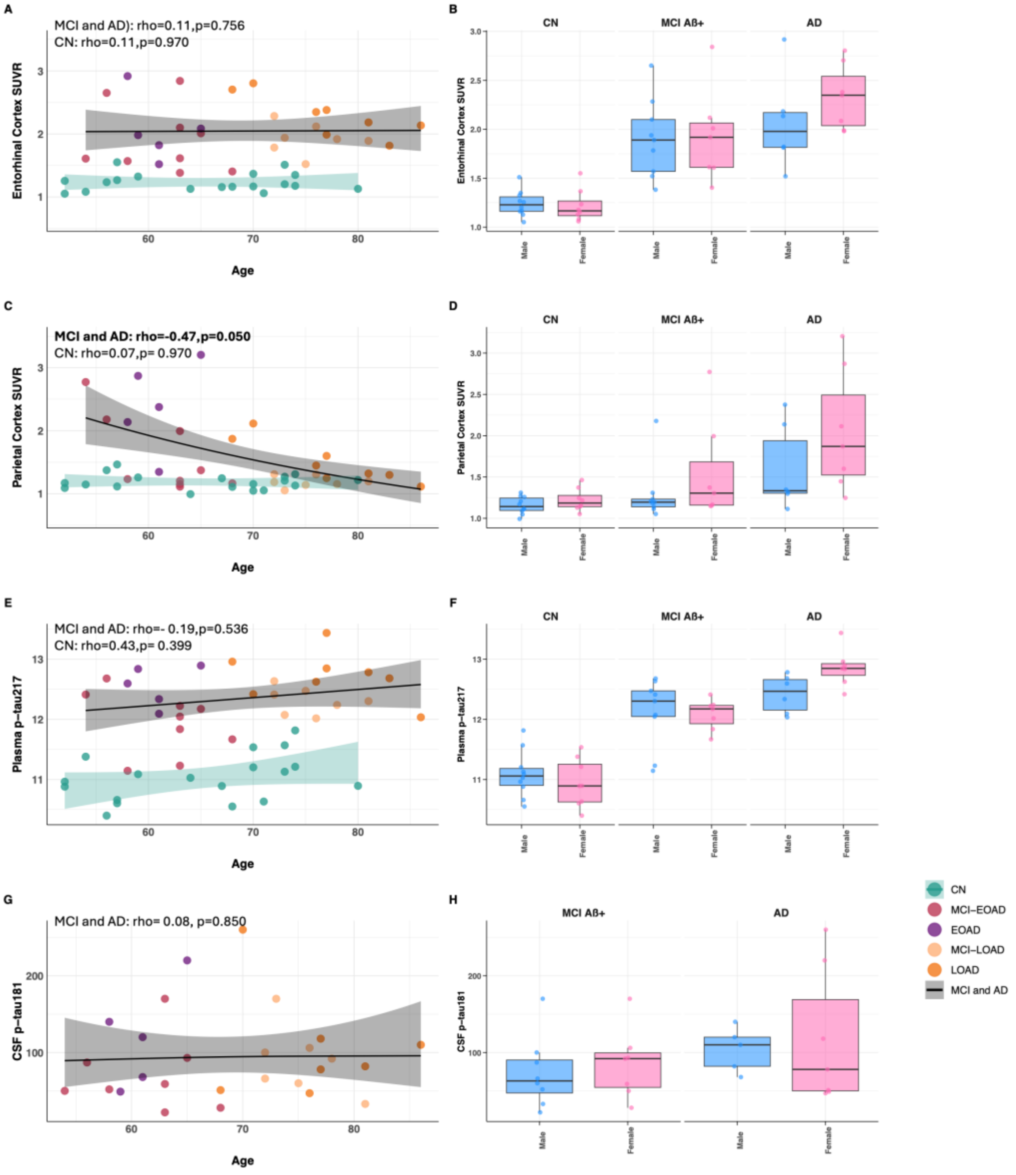


**Fig. S5. Age-related associations of [$^{18}$F]RO948 tau PET, plasma p-tau217, and CSF p-tau181, with sex differences across diagnostic groups.** Panels **(A, C)** show associations between age and tau PET binding ([$^{18}$F]RO948 SUVR) in the entorhinal cortex **(A)** and parietal cortex **(C)**. Panels **(E, G)** display associations between age and fluid biomarkers: plasma phosphorylated tau (p-tau217) **(E)** and Cerebrospinal Fluid (CSF) p-tau181 **(G)**. Spearman correlation coefficients (rho) are reported separately for amyloid-positive Mild Cognitive Impairment (MCI) and Alzheimer's Disease (AD) together, and cognitively normal (CN) groups. Smooth curves were fitted using generalized additive models (GAMs) with a Gamma distribution and log link, stratified by group. Shaded areas indicate 95% confidence intervals, and colored dots represent individual data points across five groups: CN, early-onset and late-onset with amyloid-positive MCI, and AD. FDR-corrected p-values are reported within each panel. Panels **(B, D, F, H)** show boxplots by sex for each biomarker across the diagnostic groups.

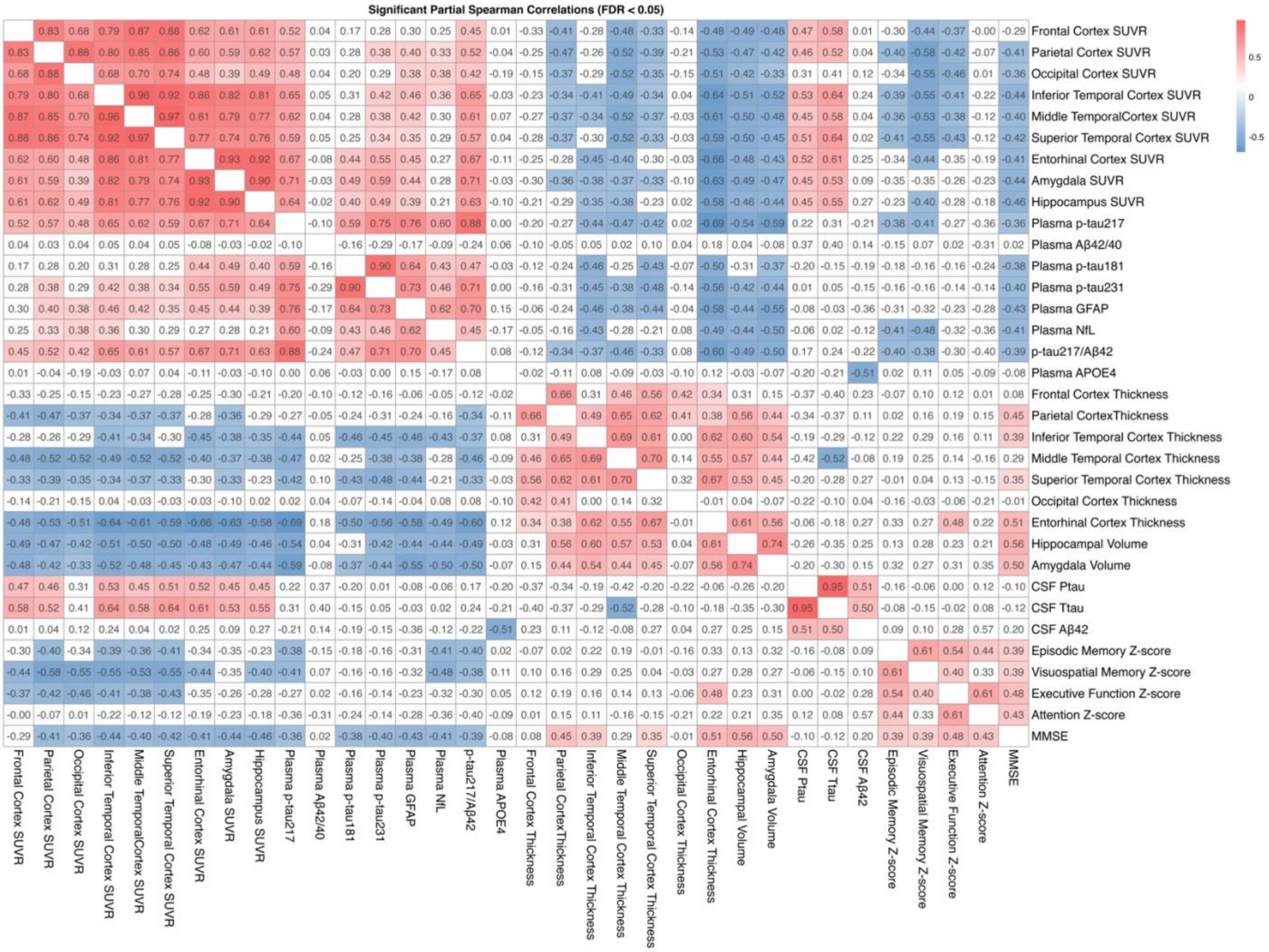


**Fig. S6. Heatmap of associations between [18F]RO948 tau PET binding, plasma biomarkers, CSF, cortical thickness and cognitive index.** Partial Spearman correlation matrix showing significant partial pairwise associations (FDR < 0.05) between regional tau PET binding ([18F]RO948), plasma biomarkers, Cerebrospinal Fluid (CSF), cortical thickness measures, and cognitive performance (MMSE, Episodic Memory Z-score, Visuospatial Memory Z-score, Executive Function Z-score, and Attention Z-score) correcting for Age, Sex and APOE status. Positive correlations are shown in red and negative correlations in blue, with intensity reflecting strength (rho value). Only statistically significant correlations (after false discovery rate correction) are displayed in color while non-significant associations are displayed in white. [18F]RO948 SUVRs, cortical thickness and plasma analyses (n=47),CSF (n=27), MMSE(n=45), Episodic Memory z-score(n=39) Visuospatial Memory Z-score (n=36), Executive Function Z-score (n=40), and Attention Z-score(n=32), Age(n=47), Education(n=32). MMSE=Mini-Mental State Examination.

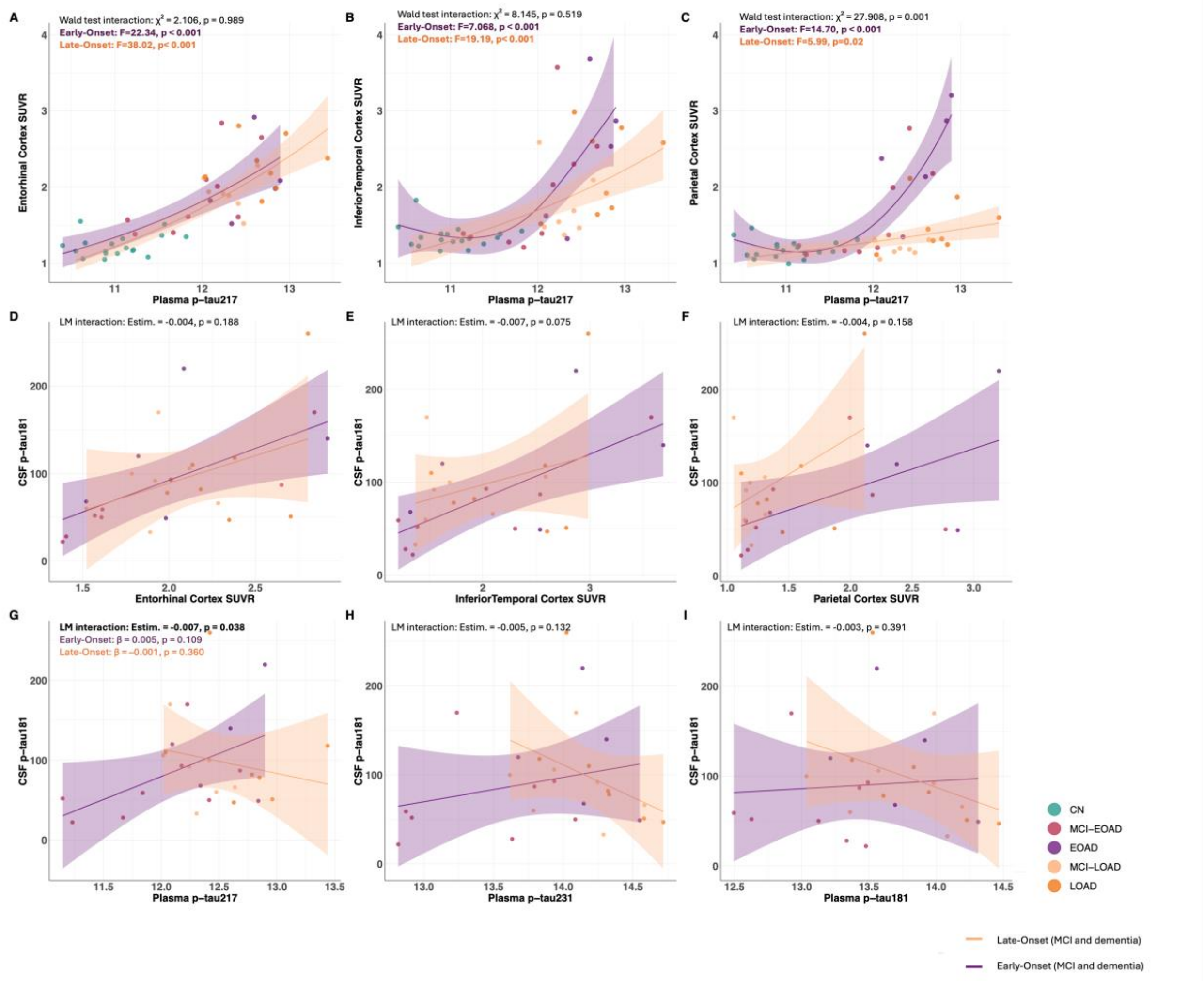


**Fig. S7. Regional regressions between [$^{18}$F]RO948 tau PET SUVRs and plasma biomarkers and their relationship with CSF p-tau181 across Alzheimer's disease diagnostic groups. (A-C)** Regressions between plasma p-tau217 and regional tau PET binding in the entorhinal **(A)**, inferior temporal cortex **(B)**, and parietal cortex **(C)**, modeled using generalized additive models (GAMs) with a gamma distribution and log-link function. Separate smooth terms are fitted for early-onset MCI and AD (purple line) and late-onset MCI and AD (orange line). Shaded areas represent 95% confidence intervals. Wald tests of the difference between the smoothed curves for EOAD and LOAD are also shown (chi-squared and p-value). **(D-I)** Associations between CSF p-tau181 and regional tau PET SUVRs in the entorhinal **(D)**, inferior temporal cortex **(E)**, and parietal cortex **(F)**, and **(G-I)** associations between CSF p-tau181 and plasma biomarkers **(G)** p-tau217, **(H)** p-tau231 and **(I)** p-tau181, modeled using linear regression models (LMs). Separate lines are fitted for early-onset AD (purple line) and late-onset AD (orange line). Shaded areas represent 95% confidence intervals. Interaction term between groups is presented with the estimate and p value. Colored dots indicate individual data points across five groups: cognitively normal (CN), early-onset and late-onset mild cognitive impairment (MCI-EOAD/MCI-LOAD) or Alzheimer's Disease (EOAD/LOAD). All analyses were controlled for Sex and APOE status as covariates. CSF= Cerebrospinal Fluid, p-tau= phosphorylated tau, SUVR= Standardized Uptake Value Ratio.

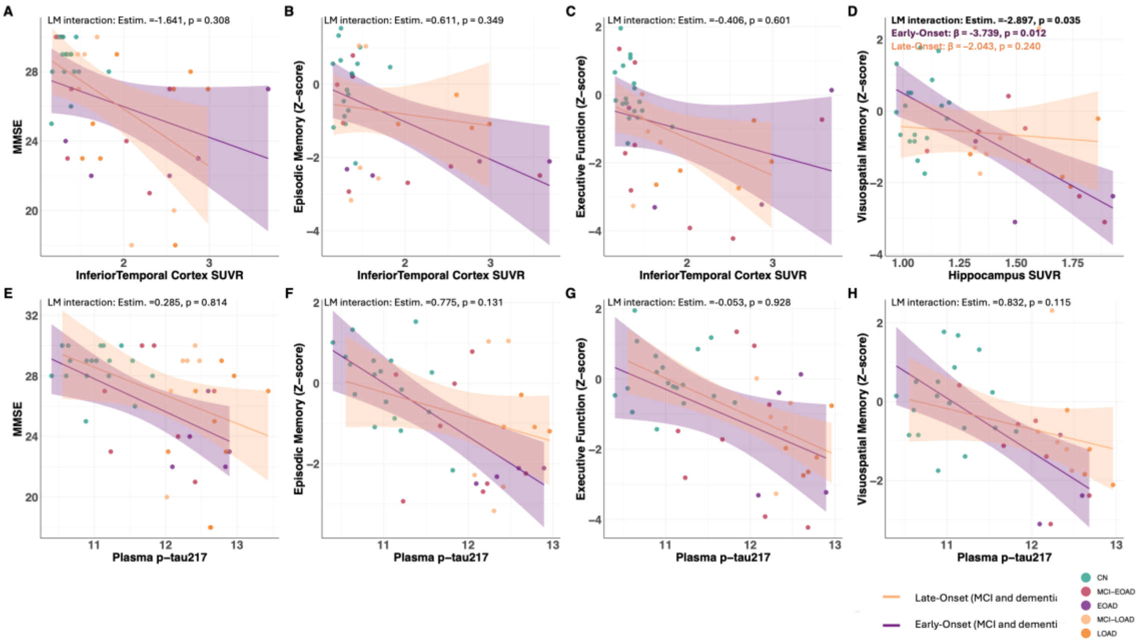


**Fig. S8. Regional regressions between [18F]RO948 tau PET binding, plasma biomarkers and cognition across Alzheimer's disease diagnostic groups. (Panels A-D)** Regressions between **(A)** Inferior Temporal [18F]RO948 SUVRs and MMSE **(B)** Inferior Temporal [18F]RO948 SUVRs and Episodic Memory **(C)**Inferior Temporal [18F]RO948 SUVRs and Executive Function **(D)** Hippocampus [18F]RO948 SUVRs and Visuospatial memory. (**Panels E-H)** Associations between plasma p-tau217 and **(E)** MMSE **(F)**Episodic Memory **(F)** Executive function and **(H)** Visuospatial memory. MMSE(n=45), Episodic Memory z-score(n=39) Visuospatial Memory Z-score (n=36), Executive Function Z-score (n=40), and Attention Z-score(n=32). Scatter plots show raw data points and unadjusted regression lines by EOAD/LOAD. Shaded areas represent 95% confidence intervals. LM interaction estimates and p-values are derived from models controlling for Sex and APOE status.p-tau= phosphorylated tau, SUVR= Standardized Uptake Value Ratio, MMSE= Mini-Mental State Examination. **Group abbreviations:** CN = cognitively normal; MCI-EOAD/MCI-LOAD= early-onset and late-onset mild cognitive impairment; EOAD/LOAD = early-onset and late-onset Alzheimer's disease.

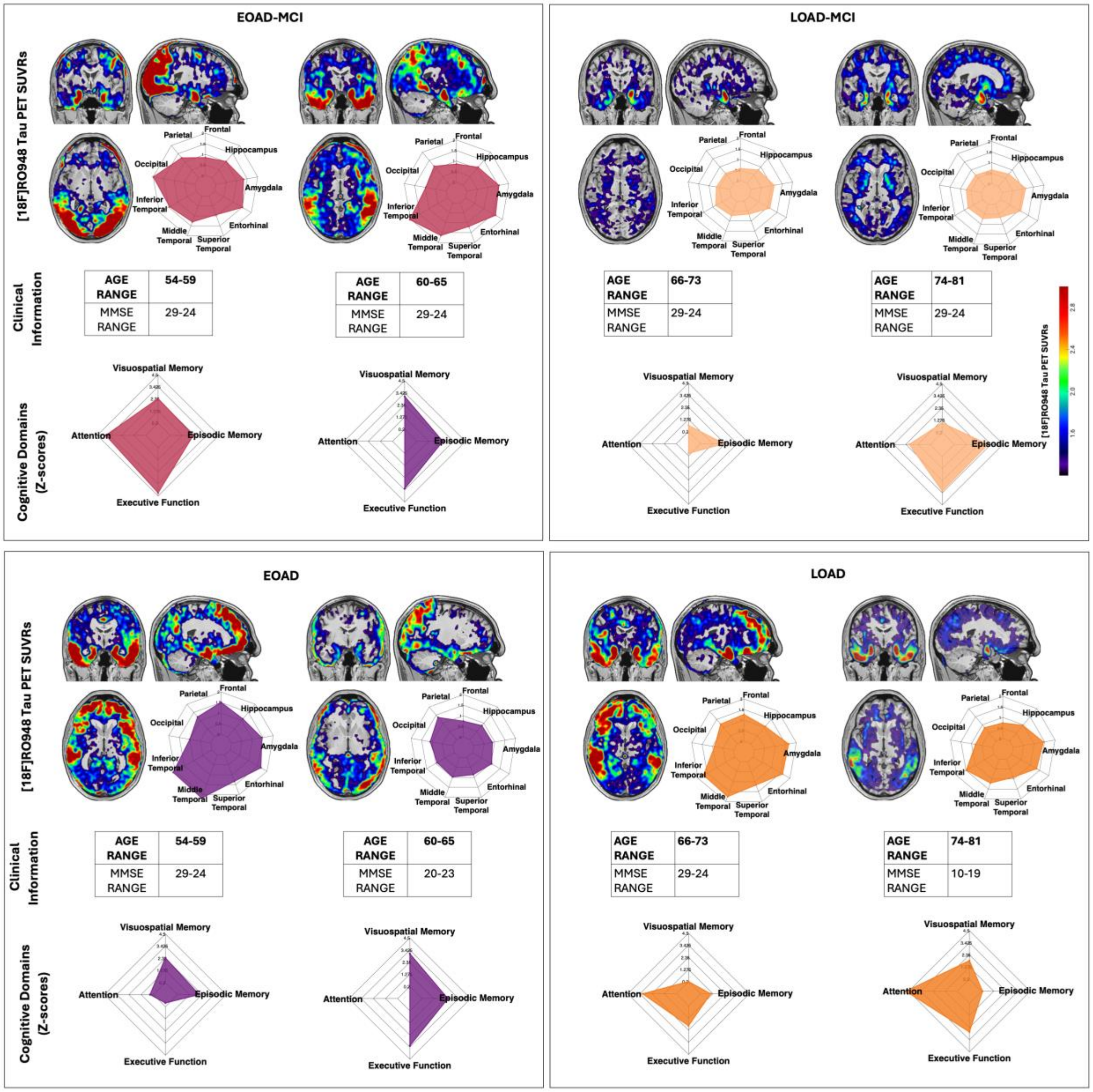


**Fig. S9. Representative [$^{18}$F]RO948 tau PET images, regional SUVR profiles, and cognitive/clinical features across Alzheimer's disease diagnostic groups.** Representative [$^{18}$F]RO948 tau-PET scans (overlayed on corresponding MRI for anatomical reference) are shown for two individuals per diagnostic group: mild cognitive impairment—early and late-onset (MCI-EOAD and MCI-LOAD), and Alzheimer's disease—early and late-onset (EOAD and LOAD). For each subject, radar plots display Standard Uptake

Value Ratios (SUVR) values across key brain regions (Frontal, Parietal, Occipital, Inferior Temporal, Middle Temporal, Superior Temporal, Entorhinal, Amygdala, Hippocampus), along with corresponding radar plots of cognitive performance profiles. Individual-level clinical data including age, years of education, and MMSE scores are also shown. Regional tau uptake increases progressively from MCI-amyloid positive (Aβ+) toward dementia stages, with distinct neocortical involvement patterns differentiating EOAD and LOAD. Radar plots emphasize group-specific topographies of tau burden and associated cognitive phenotypes. MMSE= Mini-Mental State Examination.

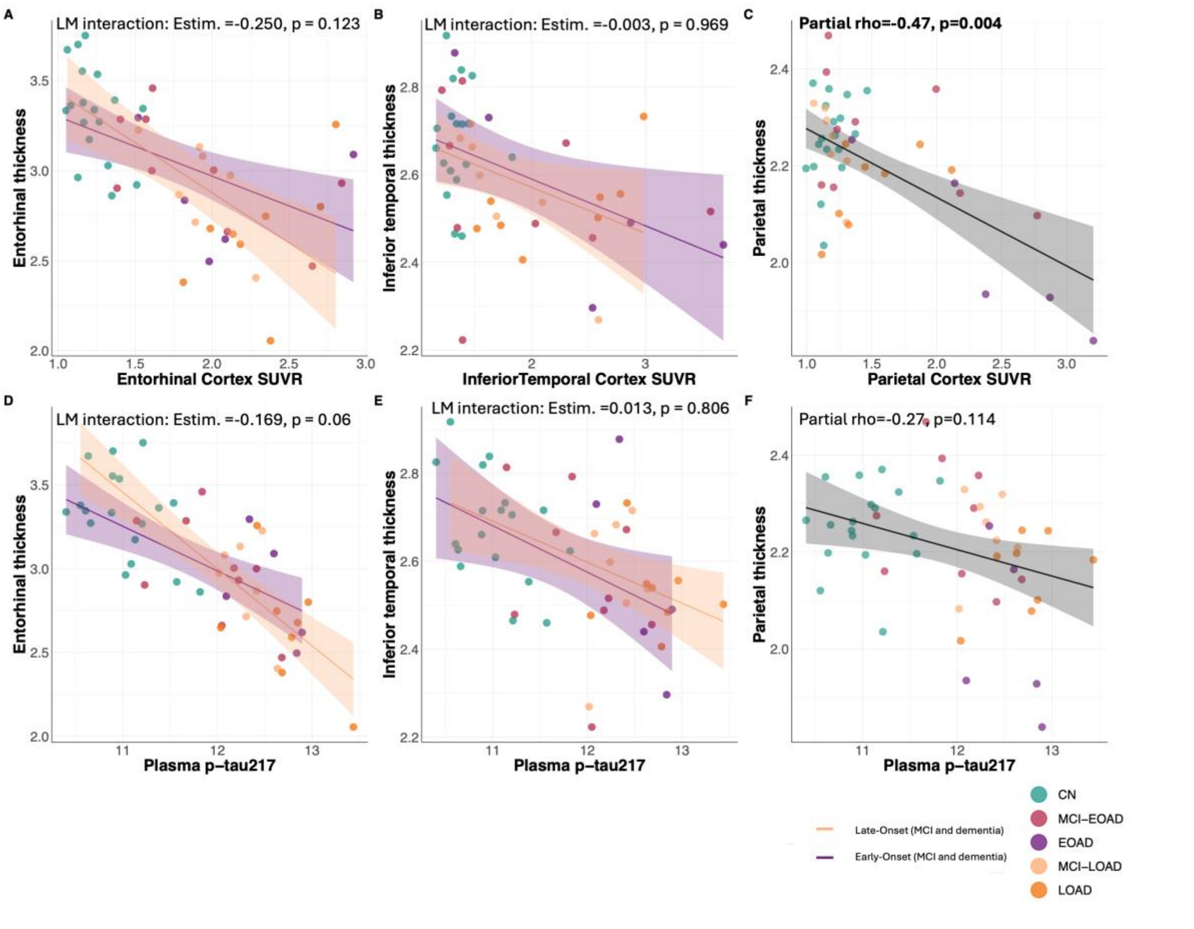


**Fig. S10. Significant correlations between Tau PET or Plasma Biomarkers and Cortical Thickness. Panels A-C)** Associations between **(A)** Entorhinal [18F]RO948 SUVR and Entorhinal thickness**(B)** Inferior Temporal [18F]RO948 SUVRs and Inferior Temporal thickness **(C)** Parietal [18F]RO948 SUVRs and Parietal thickness (**Panels D-F)** Associations between plasma p-tau217 and **(D)** Entorhinal thickness **(E)**Inferior Temporal thickness **(F)** Parietal thickness. Scatter plots show raw data points and unadjusted regression lines by EOAD/LOAD. Shaded areas represent 95% confidence intervals. LM interaction estimates and p-values are derived from models controlling for Sex and APOE status. Panel **(C,F)** fitted lines are presented for the entire sample and partial correlation coefficients (rho) and p-values are derived from models controlling for Sex and APOE status. SUVR= Standard Uptake Value Ratio. **Group**

**abbreviations:** CN = cognitively normal; MCI-EOAD/MCI-LOAD= early-onset and late-onset mild cognitive impairment; EOAD/LOAD = early-onset and late-onset Alzheimer's disease.
